# Metabolomic insight into the link of Intermuscular Fat with Cognitive Performance: The Health ABC Study

**DOI:** 10.1101/2025.01.13.25320416

**Authors:** Richard Xu, Qu Tian, Megan M. Marron, Luigi Ferrucci, Shanshan Yao, Seyoung Kim, Ravi V. Shah, Venkatesh L. Murthy, Anne B. Newman, Iva Miljkovic, Caterina Rosano

## Abstract

There is growing evidence that higher intermuscular fat (IMF) is associated with worse processing speed, measured by the digit symbol substitution test (DSST) in older adults. However, the underlying biological mechanisms are not well understood. Considering that both muscle and the brain are metabolically active organs, we sought to identify metabolites that may explain the IMF-DSST association. We assessed 613 plasma metabolites in 2388 participants from the Health, Aging and Body Composition Study (mean age ± SD: 74.7 ± 2.9 years, 50% men, 63% white), using liquid chromatography-mass spectrometry. We found that higher IMF was associated with worse DSST scores (standardized beta (95% CI): -0.08 (-0.12, -0.03), p<0.001). Sixty-six metabolites were significantly associated with both IMF and DSST. Four of the 66 metabolites attenuated the association by ≥10%: higher levels of adrenic acid (polyunsaturated fatty acid), and lower levels of C20:5 lysophosphatidylcholine (lysophospholipid), 1-methylnicotinamide (vitamin B3-related myokine), and maslinic acid (triterpene) were associated with higher IMF and worse DSST. Together, they explained 41% of the IMF-DSST association. Pathway enrichment analyses identified two significant shared pathways: unsaturated fatty acid metabolism and the citrate (TCA) cycle. This study provides hypothesis-generating evidence that a set of circulating metabolites related to unsaturated fatty acids, energy metabolism, and myokines may partially explain the inverse association of IMF with processing speed. The findings, if further confirmed by independent studies, advance our understanding of molecular pathways underlying muscle-brain crosstalk and raise the possibility of metabolites as potential predictive biomarkers and/or therapeutic targets.

## INTRODUCTION

Cognition is a crucial aspect of health among older adults, contributing significantly to independence such as managing daily activities and quality of life including social interactions and relationships. It is important to identify risk or protective factors that can affect cognition in older adults and inform targeted prevention. There is emerging evidence that cognitive function can be strongly affected by factors outside the brain, such as metabolic derangements that are associated with changes in body composition. Specifically, higher intermuscular fat (IMF) is associated with worse cognition in older adults, in particular processing speed as measured by Digit Symbol Substitution Test (DSST).^1–8^ Intermuscular fat might be a novel risk factor for cognitive decline, but the mechanisms underlying this relationship remain unclear.

IMF consists of ectopic fat depots found beneath the fascia and within the skeletal muscles.^9^ Muscle parenchyma responds to fat infiltration by releasing proinflammatory myokines (cytokines secreted by myocytes) with neurodegenerative effects.^10^ Adipose tissue stored in the muscle can also secrete proinflammatory adipokines that can affect the musculoskeletal and central nervous systems, among others. ^9^ Our hypothesis is that circulating plasma metabolites, including but not limited to those known to be related to higher IMF, may explain, at least in part, the relationship between IMF and cognitive function.

The skeletal muscle and central nervous systems share a variety of signaling pathways and molecules, including hormones, myokines, and other cytokines and neurotropic factors.^10^ Notably, skeletal muscle contractile activity (e.g. during exercise) leads to release of neuroprotective myokines.^11^ Less is known about the metabolites that relate to IMF and central nervous system. Metabolomics is a systematic approach to profile circulating metabolites;^12^ circulating metabolites are useful indicators of environment and behaviors, genetic, transcript, and protein profiles, as well as gene-environment interactions.^13^ Metabolomics may have a particular value in elucidating the communication between muscle and brain, because both tissues are metabolically active and release signaling molecules that influence metabolism and function.^14^

Here, we aimed to conduct a cross-sectional analysis of the association of IMF, circulating metabolites, and processing speed measured by the DSST in the Health, Aging and Body Composition (Health ABC) study of community-dwelling older adults. We were particularly interested in identifying metabolites that explained the association between IMF and DSST.

## METHODS

### Study Population

The Health ABC study is a prospective cohort that evaluated the impact of changes in weight and body composition on age-related physiological and functional changes among community-dwelling older Black and White men and women. The study was initiated in 1997-98 and involved 3,075 men and women who were aged 70-79 at baseline. The study included a significant representation of the Black population, with 45% of the women and 33% of the men self-reporting as Black. The study recruited individuals who were free of significant functional limitations at baseline.^15^ All the participants gave written informed consent and protocol approval at the field sites was provided by each Institutional Review Board. This analysis included a subsample of 2388 participants at the year 2 study visit who had data on intermuscular fat, metabolites, and cognitive performance (Supplemental Figure 1).

### Circulating metabolites

Overnight fasting blood samples were collected at the year 2 study visit.^16^ Plasma metabolites were measured using the following four complimentary liquid chromatography-mass spectrometry methods (LC-MS) by the Broad Institute (Cambridge, MA) between 2019-2021: (1) polar metabolite profiling method using positive ion mode MS detection, (2) polar metabolite profiling method using negative ion mode MS detection, (3) lipid profiling method, and (4) intermediate polarity profiling method. The metabolites were quantified as LC-MS peak areas, with details published elsewhere.^17,18^

A total of 613 known metabolites were detected in Health ABC plasma samples. Missing values for metabolites were assumed to be under the detectable limit and replaced with half the minimum recorded value for the respective metabolite.

### Intermuscular Fat (IMF)

Thigh computed tomography scans were collected at year 1 at the Pittsburgh site with a 9800 Advantage Scanner (General Electric, Milwaukee, WI) and either a Somatom Plus 4 (Siemens, Erlangen, Germany) or a Picker PQ 2000S (Marconi Medical Systems, Cleveland, OH) scanner at the Memphis site. ^19,20^ The scans were performed with specific technical settings (120 kVp, 200–250 mA seconds, 10 mm slice thickness) and used ILD development software (RSI Systems, Boulder, CO) for calculating tissue areas by pixel count and area. Tissues were identified based on radiographic density (HU), with calibration against distilled water (0 HU) and air (−1000 HU). Higher radiographic density indicated denser tissue with less adipose infiltration. Adipose tissue areas were specifically defined within −150 and −30 HU. The process involved distinguishing fat from lean and bone tissues in the thigh, measuring cross-sectional areas of muscle and intermuscular fat at the mid-femur level. A single 10-mm axial image of the right thigh was obtained at the midpoint of the distance between the medial edge of the greater trochanter and the intercondyloid fossa. In this image, intermuscular fat was distinguished from subcutaneous adiposity by drawing a line along the deep fascial plane surrounding the thigh muscle. The total area of non-adipose tissue and non-bone within the fascial border was quantified as the muscle thigh area. ^2^

### Cognitive outcome

Psychomotor speed was measured using the Digit Symbol Substitution Test (DSST), which tested executive function and information processing skills. It involved 9 digit-symbol pairs, and participants are required to match as many symbols to the correct digits as possible within a 90- second time frame. The DSST score (range: 0-100) is calculated based on the total number of items correctly coded, with higher scores indicating better executive functioning. The DSST was administered to participants at the baseline visit (Year 1). ^21^

### Definition of covariates

The following variables were included in the analysis: age (years), sex, and race. Total thigh muscle area was included in the analyses as it is a potential confounder for both metabolite-IMF and metabolite-DSST associations.^8^ Age (years), sex, and race were self-reported via questionnaire at baseline. ^15^

### Analytical methods

We generated summary statistics to describe the baseline characteristics of the study participants, including frequency (percent) for categorical variables and mean (standard deviation) for continuous variables. Then, we log transformed and standardized metabolites, based on their mean and standard deviation. We examined the relationship between IMF and DSST using a linear regression model adjusting for muscle area, age, sex, and race. We then conducted linear regression analyses to identify metabolites with statistically significant associations with IMF and the DSST individually, while adjusting for age, sex, race and total thigh muscle area. Benjamini-Hochberg false discovery rate (with a cutoff of FDR p<0.05) was applied to account for multiple comparisons.^22^ To identify the metabolites that significantly influenced (e.g. attenuated or strengthened) the association between IMF and DSST, we performed mediation analyses using the Mediation package (with a cutoff of FDR p<0.05); those metabolites that attenuated or strengthened the IMF-DSST association by ≥10% entered a final combined mediation model. As a complement to the individual metabolite analyses, the metabolites that were significantly associated with both IMF and DSST in these linear regression analyses were then entered into pathway enrichment analyses using MetaboAnalyst 6.0 to capture the overall patterns of metabolites significantly shared between IMF and DSST.^23^ As exploratory analyses, we also identified metabolites significantly associated with both IMF and DSST by race and sex separately and performed pathway enrichment analyses. The associations of top 4 attenuating metabolites with IMF and DSST were further analyzed to explore heterogeneity by race and sex subgroups. Analyses were conducted using R version 4.3.1.

## RESULTS

On average, the 2388 Health ABC participants were 74 years old at baseline (69-79), 50% were women, and 37% self-reported their race as Black. Age, sex, race, and thigh IMF area were significantly associated with DSST (p<0.05). Sex, race, and DSST were individually found to be significantly associated with IMF, adjusting for thigh muscle area (p<0.05, Table 1).

**Table 1.** Characteristics of 2388 Health ABC study participants.

| **Characteristics** | **Total** |  |  |
| --- | --- | --- | --- |
|  | **Frequency**<br>**(percent) or**<br>**mean (standard**<br>**deviation)** | **Associations**<br>**with DSST**<br>**beta ± standard**<br>**error** | **Associations**<br>**with IMF**<br>**(adjusted for**<br>**muscle area)**<br>**beta ± standard**<br>**error** |
| **Age (year 2), years** | 74.7 (2.9) | -0.69 (0.10) | -0.10 (0.09) |
| **Women** | 1195 (50.0) | 3.48 (0.58) | 8.86 (0.70) |
| **Race** |  |  |  |
| White | 1505 (63.0) | Ref. | Ref. |
| Black | 883 (37.0) | -13.21 (0.55) | 5.22 (0.53) |
| **Thigh intermuscular fat**<br>**(IMF) area (year 1), cm<sup>2</sup>** | 20.3 (12.8) | -0.18 (0.02) | -- |
| **Thigh muscle area**<br>**(year 1), cm<sup>2</sup>** | 223.6 (55.6) | -0.04 (0.01) | -- |
| **Digit Symbol**<br>**Substitution Test**<br>**(DSST, year 1)** | 36.4 (14.4) | -- | -0.12 (0.02) |

Associations between intermuscular fat and cognitive performance were statistically significant. Both smoothing plots (Figure 1) and linear regression analyses showed an inverse association between thigh IMF area and the DSST (beta (95% CI): -0.08 (-0.12, -0.03), p<0.001, Supplemental Table 1), such that higher IMF values were associated with worse DSST scores.

**Figure 1.**
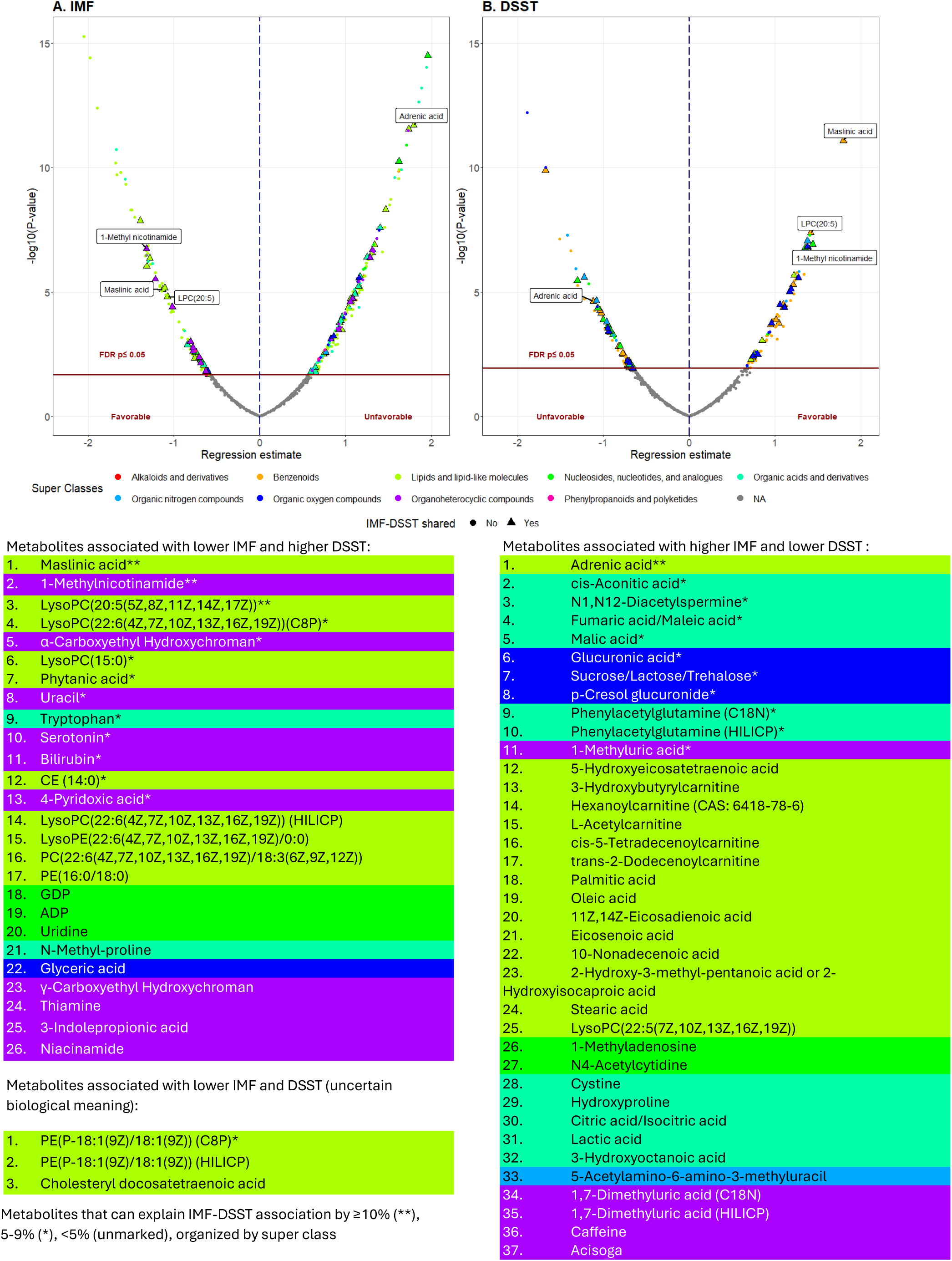
Smoothing plot of association between intermuscular fat (IMF) with digit symbol substitution test (DSST). Shaded area indicates 95% confidence interval.

Of the 613 metabolites included in this analyses, 269 metabolites were associated with IMF with FDR<0.05, and 150 metabolites were associated with DSST with FDR<0.05, both adjusting for age, sex, race as well as muscle area. A total of 66 metabolites were significantly associated with both IMF and DSST with FDR<0.05 (Table 2, Figure 2, Supplemental Table 2). For 37 of these 66 metabolites, higher metabolite values were associated with higher IMF and lower DSST scores. These 37 metabolites are classified as lipids or lipid-like molecules (n=15); organic acids and derivatives (n=11); organoheterocyclic compounds (n=5); organic oxygen compounds (n=3); nucleosides, nucleotides, and analogues (n=2), or organic nitrogen compounds (n=1). Another twenty-six of the 66 metabolites had the opposite association with IMF and DSST: higher values were associated with less IMF and better DSST scores. These 26 metabolites were classified as lipids or lipid-like molecules (n=10); organoheterocyclic compounds (n=10); nucleosides, nucleotides, and analogues (n=3); organic acids and derivatives (n=2); or organic oxygen compounds (n=1) (Figure 2). Higher levels of the remaining 3 metabolites, all classified as lipids and lipid-like molecules, were associated with lower IMF, but worse DSST.

**Figure 2.**
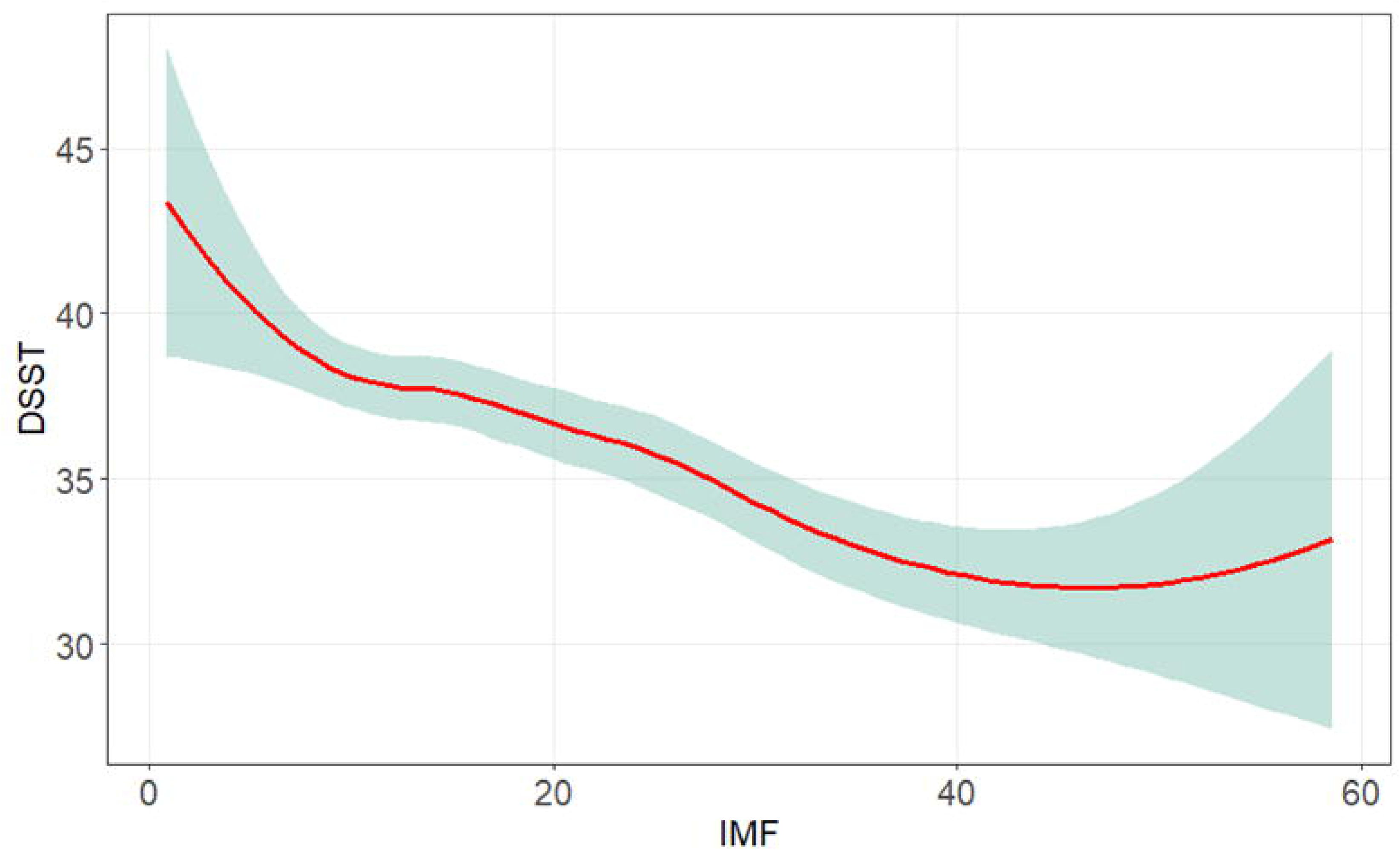
Graphic abstract: top panels: volcano plots for the associations between circulating metabolites and intermuscular fat (IMF, Panel A) and digit symbol substitution test (DSST, Panel B); bottom panels: list of significant metabolites associated with lower IMF and higher DSST, metabolites associated with higher IMF and lower DSST, and undetermined, shared metabolites between IMF and DSST in Health ABC.

**Table 2.** Metabolites with statistically significant associations with both intermuscular fat (IMF) and digit symbol substitution test (DSST), organized by class.

| Metabolites inversely associated with IMF and positively associated with DSST |  |  |  |  |  |  |  |
| --- | --- | --- | --- | --- | --- | --- | --- |
|  |  | IMF |  |  | DSST |  |  |
| Superclass/ Metabolites | Class/ Subclass | Beta | 95% CI | FDR P | Beta | 95% CI | FDR P |
| **Lipids and lipid-like molecules** |  |  |  |  |  |  |  |
| LysoPC(22:6(4Z,7Z,10Z,13Z,16Z,19Z)) (C8P) | Glycerophospholipids/<br>*Glycerophosphocholines* | -1.39 | (-1.87, -0.91) | 2.75E-07 | 0.95 | (0.45, 1.45) | 2.20E-03 |
| LysoPC(20:5(5Z,8Z,11Z,14Z,17Z)) | Glycerophospholipids/<br>*Glycerophosphocholines* | -1.08 | (-1.56, -0.59) | 9.87E-05 | 1.41 | (0.91, 1.92) | 4.53E-06 |
| LysoPC(15:0) | Glycerophospholipids /<br>*Glycerophosphocholines* | -1.32 | (-1.84, -0.79) | 9.77E-06 | 1.04 | (0.50, 1.59) | 1.90E-03 |
| Phytanic acid | Prenol lipids / *Diterpenoids* | -1.28 | (-1.78, -0.79) | 5.27E-06 | 1.01 | (0.49, 1.53) | 1.48E-03 |
| Maslinic acid | Prenol lipids/ *Triterpenoids* | -1.13 | (-1.63, -0.64) | 5.77E-05 | 1.80 | (1.28, 2.31) | 2.48E-09 |
| CE (14:0) | Steroids and steroid derivatives /<br>*Steroid esters* | -0.73 | (-1.23, -0.23) | 0.01 | 1.10 | (0.58, 1.62) | 5.17E-04 |
| **Organic acids and derivatives** |  |  |  |  |  |  |  |
| Tryptophan | Carboxylic acids and derivatives/<br>*Amino acids, peptides, and analogues* | -0.84 | (-1.36, -0.33) | 4.94E-03 | 1.44 | (0.91, 1.97) | 7.33E-06 |
| **Organoheterocyclic compounds** |  |  |  |  |  |  |  |
| α-Carboxyethyl Hydroxychroman | Benzopyrans / *1-benzopyrans* | -1.22 | (-1.73, -0.71) | 2.68E-05 | 1.27 | (0.74, 1.80) | 7.41E-05 |
| Uracil | Diazines / *Pyrimidines and pyrimidine derivatives* | -0.78 | (-1.27, -0.29) | 6.16E-03 | 1.29 | (0.78, 1.79) | 2.40E-05 |
| Serotonin | Indoles and derivatives / *Tryptamines and derivatives* | -0.81 | (-1.29, -0.33) | 3.77E-03 | 1.06 | (0.56, 1.56) | 5.17E-04 |
| 4-Pyridoxic acid | Pyridines and derivatives /<br>*Pyridinecarboxylic acids and derivatives* | -0.69 | (-1.19, -0.19) | 0.02 | 1.17 | (0.65, 1.69) | 2.05E-04 |
| 1-Methylnicotinamide | Pyridines and derivatives /<br>*Pyridinecarboxylic acids and derivatives* | -1.32 | (-1.82, -0.83) | 2.89E-06 | 1.38 | (0.87, 1.89) | 8.47E-06 |
| Bilirubin | Tetrapyrroles and derivatives/ *Bilirubins* | -0.78 | (-1.28, -0.28) | 7.99E-03 | 1.19 | (0.67, 1.71) | 1.65E-04 |

**Metabolites positively associated with IMF and inversely associated with DSST**
|  |  | IMF |  |  | DSST |  |  |
| --- | --- | --- | --- | --- | --- | --- | --- |
| Superclass/ Metabolites | Class/ Subclass | Beta | 95% CI | FDR P | Beta | 95% CI | FDR P |
| **Lipids and lipid-like molecules** |  |  |  |  |  |  |  |
| Adrenic acid | Fatty Acyls / Fatty acids and conjugates | 1.79 | (1.30, 2.29) | 1.48E-10 | -1.12 | (-1.64, -0.60) | 4.07E-04 |
| **Organic acids and derivatives** |  |  |  |  |  |  |  |
| N1,N12-Diacetylspermine | Carboximidic acids and derivatives / Carboximidic acids | 1.15 | (0.65, 1.65) | 4.74E-05 | -1.07 | (-1.58, -0.55) | 6.13E-04 |
| Phenylacetylglutamine (C18N) | Carboxylic acids and derivatives/ Amino acids, peptides, and analogues | 0.96 | (0.48, 1.44) | 4.98E-04 | -0.89 | (-1.39, -0.39) | 3.71E-03 |
| Phenylacetylglutamine (HILICP) | Carboxylic acids and derivatives/ Amino acids, peptides, and analogues | 0.94 | (0.46, 1.43) | 6.48E-04 | -0.95 | (-1.45, -0.44) | 2.22E-03 |
| Fumaric acid/Maleic acid | Carboxylic acids and derivatives / Dicarboxylic acids and derivatives | 1.11 | (0.61, 1.60) | 8.20E-05 | -0.95 | (-1.46, -0.44) | 2.62E-03 |
| cis-Aconitic acid | Carboxylic acids and derivatives / Tricarboxylic acids and derivatives | 1.40 | (0.91, 1.89) | 4.83E-07 | -0.92 | (-1.43, -0.41) | 3.31E-03 |
| Malic acid | Hydroxy acids and derivatives / Beta hydroxy acids and derivatives | 1.09 | (0.59, 1.58) | 1.13E-04 | -1.00 | (-1.52, -0.49) | 1.48E-03 |
| **Organic oxygen compounds** |  |  |  |  |  |  |  |
| Glucuronic acid | Organooxygen compounds / Carbohydrates and carbohydrate conjugates | 1.16 | (0.67, 1.64) | 2.45E-05 | -0.96 | (-1.47, -0.46) | 1.77E-03 |
| Sucrose/Lactose/Trehalose | Organooxygen compounds / Carbohydrates and carbohydrate conjugates | 0.86 | (0.37, 1.36) | 2.47E-03 | -1.22 | (-1.73, -0.72) | 7.41E-05 |
| p-Cresol glucuronide | Organooxygen compounds / Carbohydrates and carbohydrate conjugates | 0.83 | (0.35, 1.31) | 3.06E-03 | -1.08 | (-1.58, -0.58) | 4.05E-04 |
| **Organoheterocyclic compounds** |  |  |  |  |  |  |  |
| 1-Methyluric acid | Imidazopyrimidines/ Purines and purine derivatives | 0.92 | (0.42, 1.42) | 1.41E-03 | -0.94 | (-1.46, -0.42) | 3.31E-03 |

In mediation analyses applied to the 66 metabolites that were associated with both IMF and DSST, we found 4 metabolites attenuated the IMF-DSST association by ≥10% when examined individually. Higher levels of one metabolite, adrenic acid (12%, fatty acyls), was associated with more IMF and worse DSST scores. Whereas higher levels of the other three were associated with less IMF and better DSST scores: C20:5 lysophosphatidylcholine (10% attenuation, lysophospholipid containing omega-3 fatty acid), 1-methylnicotinamide (12% attenuation, myokine derived from vitamin B3), and maslinic acid (14% attenuation, triterpene found in olives). We also found 21 metabolites attenuated the IMF-DSST association by >5% and <10% (Table 3, Figure 3), including lipids or lipid-like molecules (n=5), organic acids and derivatives (n=7), organoheterocyclic compounds (n=6), or organic oxygen compounds (n=3). These metabolites were not included in further mediation analyses.

**Figure 3.**
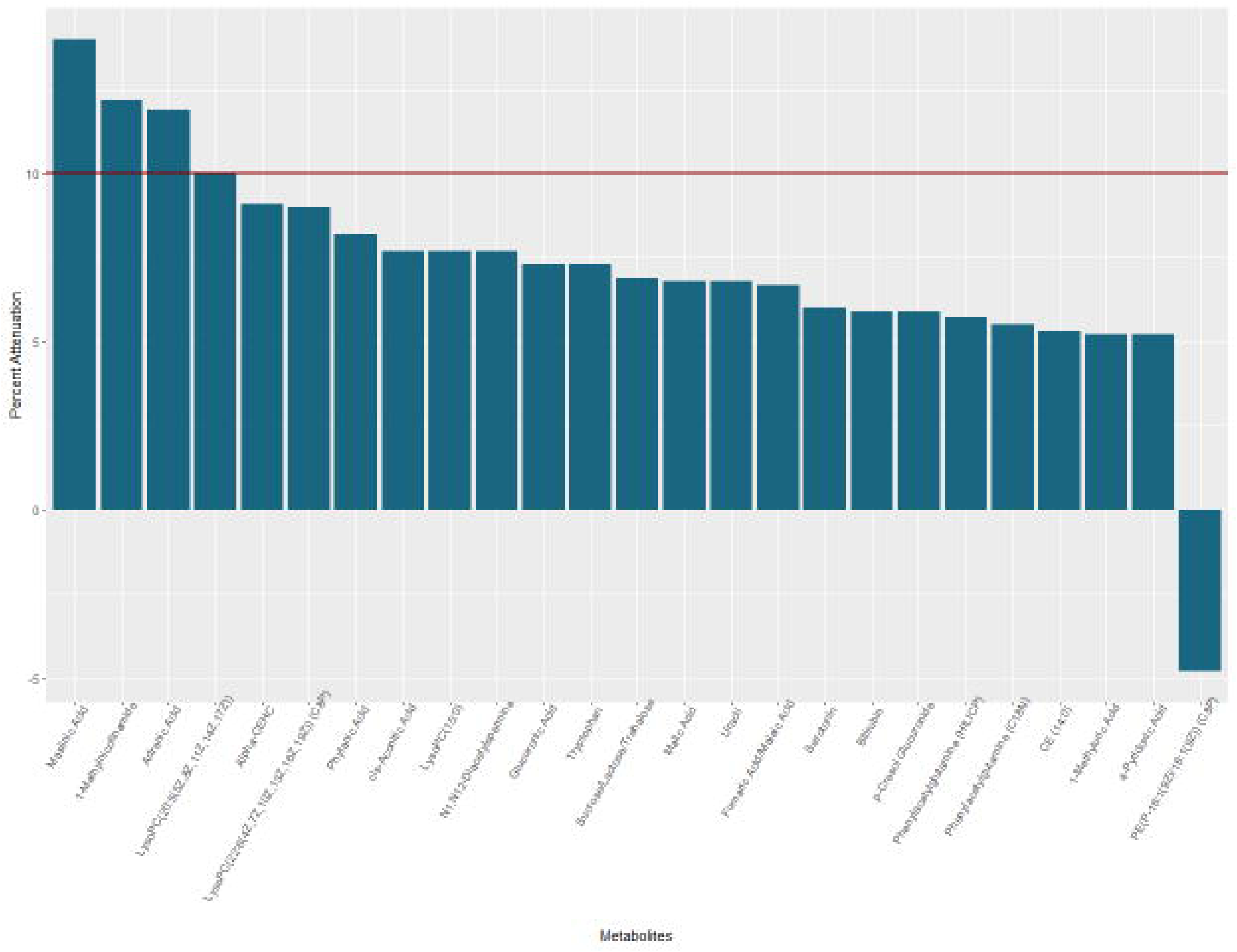
Percent attenuation or enhancement in the association between intermuscular fat and digit symbol substitution test among top 25 significantly attenuating and strengthening metabolites

**Table 3.**
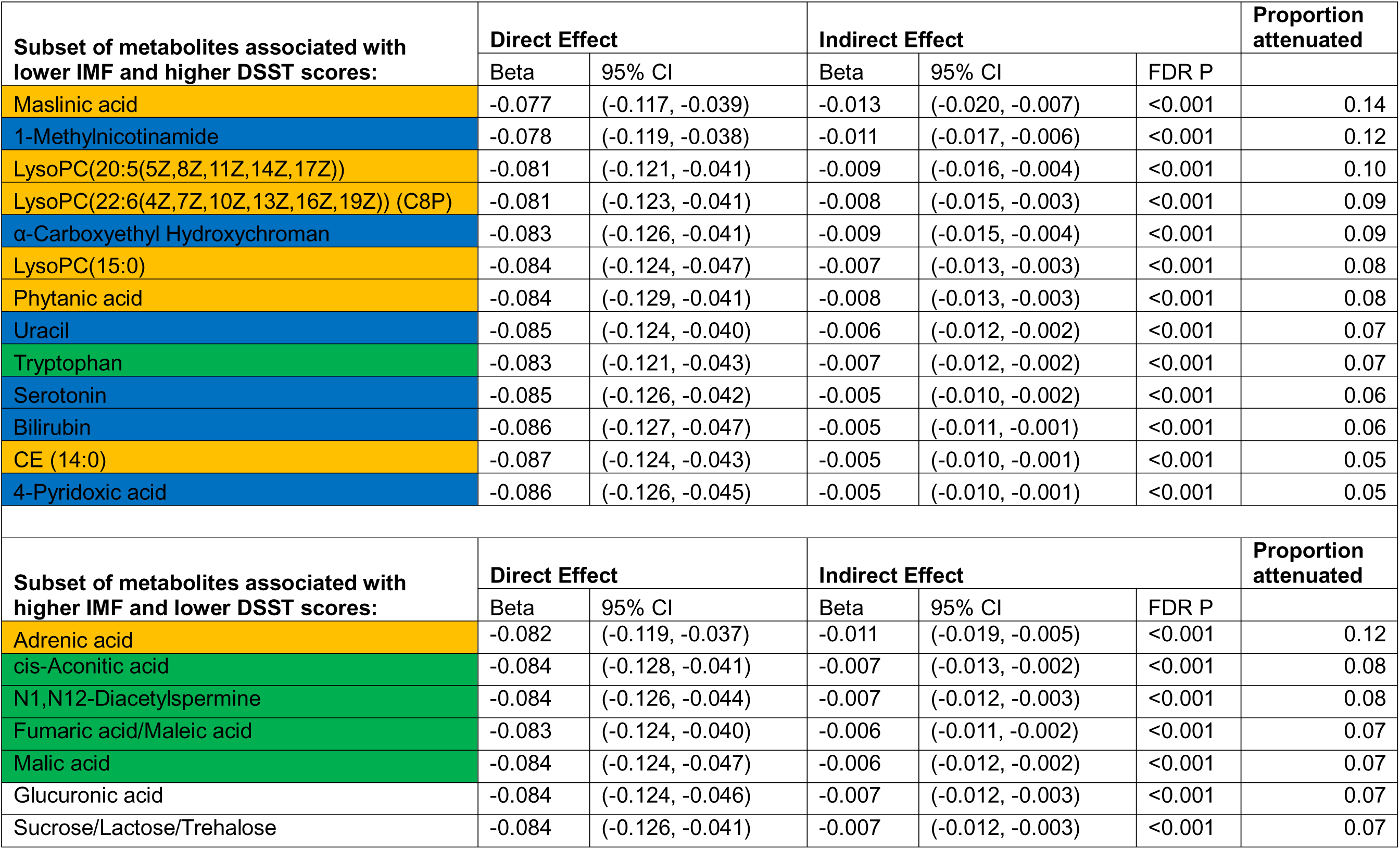
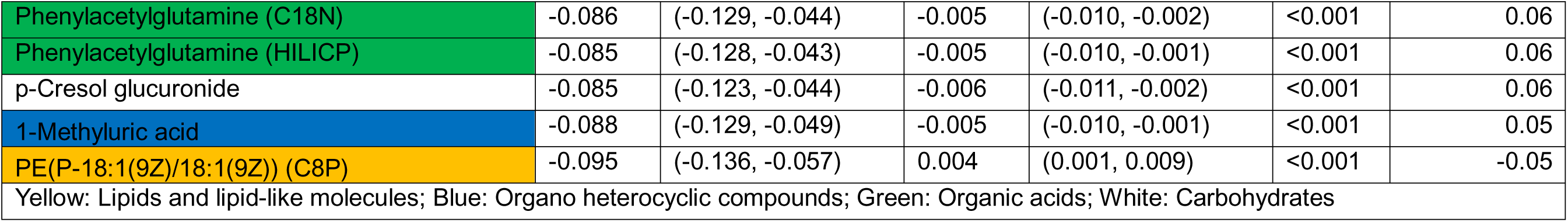
Metabolites that are significantly associated with intermuscular fat (IMF) and the digit symbol substitution test (DSST) and also significantly enhancing or attenuating the association between IMF and DSST.

A final combined mediation analysis included the 4 top mediating metabolites. The total effect of IMF on DSST is -0.091, meaning that for every 1 cm2 increase in IMF, DSST decreases by 0.091 points. 41% of this association is attenuated through the top 4 metabolites: maslinic acid (11%), 1-MNA (11%), LPC(20:5) (7%), and adrenic acid (11%), indicating that these 4 metabolites together attenuate a significant portion of the association between IMF and DSST (Supplemental Table 3).

Functional annotations for metabolites that attenuated or strengthened the IMF-DSST association by at least 5% are provided in Supplemental Table 4 and Supplemental Table 5.

As a complementary analysis to the individual metabolites, we performed pathway enrichment analysis on the 66 metabolites significantly associated with both IMF and DSST. We found two shared pathways that contained significantly more metabolites than expected for the pathways by chance. One was the citrate cycle (TCA cycle, FDR p=0.02067), with four shared metabolites between IMF and DSST including malic acid, cis-aconitic acid, citric acid, and fumaric acid. The other pathway was the biosynthesis process of unsaturated fatty acids (FDR p=0.00371) with three shared metabolites between IMF and DSST including palmitic acid, stearic acid, and oleic acid (Supplemental Figure 2).

Finally, we performed subgroup analyses by race and sex separately. We first explored whether the associations between the top 4 metabolites with IMF and DSST were consistent across race and sex subgroups as shown in Supplemental Figure 3. In general, the patterns were consistent across the subgroups; however, black individuals had higher IMF and lower DSST for any given metabolite level compared to white individuals. Within a racial group, there were no clear differences in IMF, but females had higher DSST for a given metabolite value.

We also analyzed race- or sex-specific metabolomic association with IMF and DSST. Among White participants, 78 metabolites were significantly associated with both IMF and DSST, of which adrenic acid; LysoPC(20:5); LysoPC(22:6); maslinic acid; CE(14:0); phenylacetylglutamine (C18N); phenylacetylglutamine (HILICp); tryptophan; N1,N12- diacetylspermine; fumaric acid/maleic acid; cis-aconitic acid; malic acid, p-cresol glucuronide; sucrose/lactose/trehalose; glucuronic acid; uracil; 1-methyluric acid; 1-MNA; and bilirubin were consistent with the total sample (Supplemental Table 6). Among Black participants, 5 metabolites were significantly associated with both IMF and DSST, of which LysoPC(15:0), 1- MNA, and serotonin, were consistent with the total sample (Supplemental Table 7). Among male participants, 24 metabolites were significantly associated with both IMF and DSST, of which adrenic acid, LysoPC(20:5), LysoPC(22:6), phytanic acid, maslinic acid, phenylacetylglutamine (C18N); phenylacetylglutamine (HILICp), tryptophan, fumaric acid/maleic acid, malic acid, p-cresol glucuronide, sucrose/lactose/trehalose, glucuronic acid, 1-MNA, and bilirubin were consistent with the total sample (Supplemental Table 8). Among female participants, 9 metabolites were significantly associated with both IMF and DSST, of which maslinic acid; N1,N12-diacetylspermine, α-Carboxyethyl Hydroxychroman, and 1-MNA were consistent with the total sample (Supplemental Table 9). For White participants, pathway enrichment analyses identified biological pathways consistent with the total sample: citrate cycle (TCA cycle) and biosynthesis of unsaturated fatty acids in addition to glyoxylate and dicarboxylate metabolism (Supplemental Figure 4). No significant biological pathways were identified for Black, male, or female participants separately. Insufficient metabolites were identified for lipid biological pathways (Supplemental Figures 5,6,7). We were unable to perform subgroup analyses by both race and sex simultaneously due to limited sample size.

## DISCUSSION

To our knowledge, this is the first study in a cohort of community dwelling older adults, to explore metabolomic pathways underlying the relationship of IMF and processing speed. We identified 66 metabolites shared among IMF and DSST, related to unsaturated fatty acids and energy metabolism; of these, 4 metabolites together explained 41% of the IMF-DSST association.

Our results are consistent with prior metabolomic studies in older age in relation with indices of brain and muscle health, unlike prior studies that assessed brain and muscle health in separate models, our study present a novel approach by assessing both brain and muscle health indices in the same model. A systematic review by Tian et al. found five classes of metabolites, including fatty acids, phosphatidylcholines, and biogenic amines, that were shared between worse cognitive function and slower gait, a proxy of muscle health.^24^ Another recent study found plasma lysoPCs and other lipids (including glycerophospholipids, sphingolipids) were associated with mobility decline and dementia risk.^25^ Our findings add to this body of evidence, by examining a direct marker of muscle health, IMF, which is emerging as a novel risk factor for cognitive impairment in older adults.

Pathway enrichment analysis indicated two shared pathways between IMF and DSST: biosynthesis of unsaturated fatty acids and citrate cycle (TCA cycle). Dysfunction in the TCA cycle could result in altered energy metabolism and oxidative stress that could affect both muscle and the brain. Unsaturated fatty acids maintain and regulate tissue function through their roles in energy production. For example, omega-3 and omega-6 poly-unsaturated fatty acids maintain membrane fluidity, which is crucial for muscle contractility^26^ and synaptic remodeling. ^27^ Some omega-3 polyunsaturated fatty acids have potent anti-inflammatory properties.^28^ Omega- 3 fatty acids have been shown to enhance muscle protein synthesis, which can help prevent sarcopenia in older adults,^29^ and to have neuroprotective effects, reducing oxidative stress and inflammation in Alzheimer’s and Parkinson’s disease. ^30^

Adrenic acid, an omega-6 polyunsaturated fatty acid (22:4n-6 or 7,10,13,16-docosatetraenoic acid), was among the top 4 metabolites that explained the IMF-DSST association by ≥10%. Specifically, we found higher levels of adrenic acid were correlated with higher IMF and worse DSST. Although a pro-inflammatory role of adrenic acid has been shown, a recent study suggested a neuroprotective effect of adrenic acid, in particular with regard to brain Aβ load.^31^ Adrenic acid may have hormetic adaptive responses in relation to inflammatory processes; similar to other myokines, such as interleukin-6 which has anti-inflammatory effects after muscle contractions and pro-inflammatory effects in obese individuals.^32,33^ The diverse and bi- directional functions of adrenic acid with muscle and brain health underline the complexity and significance of omega-6 fatty acids in human health, challenging the simplistic classification of omega-6 fatty acids as solely pro-inflammatory. Further research is needed to fully elucidate its functions and mechanisms of action in various physiological contexts.

Another metabolite related to fatty acids, C20:5 lysophosphatidylcholine (LysoPC(20:5)), was also among the 4 metabolites explaining the IMF-DSST association by ≥10%. Higher levels of LysoPC were associated with lower IMF and higher DSST, suggesting this metabolite may be linked to favorable health profiles of muscle and brain. This is consistent with a prior study showing higher LysoPC(20:3) concentration was associated with higher skeletal muscle mitochondrial oxidative capacity.^34^ In a study of patients with mild to moderate AD, higher LysoPC appeared to play a role in maintaining cognitive function.^35^ Similarly, in a preclinical study, lower levels of LysoPC were associated with greater skeletal muscle myopathy. ^36^ Of note, a small case control study from the Baltimore Longitudinal Study on Aging^37^ found higher levels of LysoPC were related with lower muscle quality; however, in that study muscle quality was operationalized as the ratio between mass and force, which is only slightly related to IMF. Further studies of LysoPC(20:5) are warranted to better understand IMF-cognition associations.

Similar to LysoPC, the myokine 1-MNA (1-methylnicotinamide) was higher for those with lower IMF and better DSST and was among the 4 metabolites explaining the IMF-DSST association. 1-MNA, a derivative of niacin (vitamin B3), is emerging as a novel myokine that enhances the utilization of energy stores in response to low muscle energy availability. ^38^ 1-MNA can function through both intracellular and extracellular means. ^8^ Plasma levels of 1-MNA increased after a high-volume-low-intensity exercise intervention combined with caloric restriction in men between the ages of 18-55. This increase in 1-MNA was mirrored by increased expression of Nicotinamide N-methyltransferase in skeletal muscle, the enzyme involved in producing MNA. ^38^ Neuroprotective effects of 1-MNA have recently been reported in animal models of inflammation and Alzheimer’s. Higher plasma levels of 1-MNA significantly attenuated lipopolysaccharide-induced cognitive deficits in 3-month-old mice, possibly by inhibiting the activation of glial cells and NF-κB signaling. ^39^ In another mouse study, administration of 1-MNA significantly reversed cognitive impairments after bilateral intrahippocampal injection of Aβ1-42. Furthermore, 1-MNA suppressed Aβ1-42-induced neuroinflammation, characterized by suppressed activation of microglia, decreased the expression of IL-6, TNF-α and nuclear translocation of NF-κB p65, as well as attenuated neuronal apoptosis. ^40^ Taken together, these data indicate 1-MNA may be a promising myokine with neuroprotective effects, potentially linking favorable muscle and brain health profiles, and merits further investigation, especially in human prospective cohort studies or clinical trials.

The myoprotective effects of maslinic acid have also been reported. A randomized, double-blind, placebo-controlled trial of 69 healthy Japanese adult men and women found beneficial effects of maslinic acid intake on grip strength and trunk muscle mass. ^41^ Another parallel, double-blind, randomized, placebo-controlled trial found beneficial effects of maslinic acid supplementation on skeletal muscle mass, segmental muscle mass and knee pain score. ^42^ In addition to myoprotective effects, animal models and in vitro cell lines suggest maslinic acid may have neuroprotective effects through oxidative stress or inflammation processes. ^43^ In cultured cortical astrocytes, maslinic acid appeared to reduce neuroinflammation by inhibiting NF-κB signal transducer pathway. ^44^ Maslinic acid also diminished Kynurenin Acid-induced increase in hippocampal cyclooxygenase-2 activity and relative NF-κB p50/65 binding activity.^45^ Most studies assessing the effects of maslinic acid have been conducted in animal models and cell lines; future studies should assess its potential use in humans.

How should we interpret the mediation effects of these metabolites? It is possible that circulating metabolites may act as messengers of muscle-brain communication; our study points to metabolites related to unsaturated fatty acid metabolism and energy metabolism as potential messengers of muscle-brain cross-talk Skeletal muscles are highly active in energy production through various metabolic pathways, and through the release of various signaling molecules, including myokines, they regulate metabolism, inflammation, and overall metabolic health.^10^ Future studies should assess whether muscle-driven metabolic pathways also influence brain health in older age and the underlying mechanisms.

Our results should be interpreted with the constraints of a cross-sectional design. We were unable to delineate temporal relationships required to truly assess mediation. Although we performed subgroup analyses by race and sex separately, we do not have adequate sample size for analyses by race and sex simultaneously. While the overall patterns of the associations appear to be consistent between White participants and the total sample, future studies with larger sample sizes are needed to further explore other subgroups. Future studies should also consider whether medication use could alter the association between metabolites, IMF, and DSST. Finally, although the study population was community-based, caution is needed to generalize the findings to other populations with different socio-demographic characteristics.

**In conclusion,** in a sample of the Health ABC study, we identified 66 circulating metabolites that were associated with and shared between IMF and the DSST related to fatty acid and energy metabolic pathways. Four of these metabolites together attenuated 41% of the IMF- DSST association. Additional studies assessing their role in the shared pathophysiology of intermuscular fat and cognitive decline in older adults are warranted.

## Data Availability

Researchers may request Health ABC data from the NIA website at https://www.nia.nih.gov/healthabc-study. Analytical methods and R code for this study are available from corresponding author upon reasonable request.

## Acknowledgements

The authors wish to thank the study participants in the Health ABC study and the field team for their contributions to the Health ABC cohort.

## Author Contributions

Caterina Rosano, Qu Tian, Iva Miljković, Luigi Ferrucci, Ravi V. Shah, Venkatesh L. Murthy, and Anne B. Newman contributed to the study conception and design. Material preparation and data collection was performed by Caterina Rosano, Qu Tian, Iva Miljković, Luigi Ferrucci, Ravi V. Shah, Venkatesh L. Murthy, Anne B. Newman, Megan M. Marron, and Shanshan Yao. Analysis and data presentation were performed by Qu Tian and Richard Xu with input from Caterina Rosano, Megan M. Marron, and Shanshan Yao. The first draft of the manuscript was written by Richard Xu and all authors commented on previous versions of the manuscript. All authors read and approved the final manuscript. Funding was acquired by Ravi V. Shah, Venkatesh L. Murthy, and Anne B. Newman. This work is supervised by Caterina Rosano.

## Abbreviations

Health ABC: Health, Aging and Body Composition Study IMF: Intermuscular fat DSST: Digit symbol substitution test

