## Supplemental Files for "Metabolomic insight into the link of Intermuscular Fat with Cognitive Performance: The Health ABC Study"

**Supplementary Material (for online data supplement purposes)**

**Supplemental Table 1. Linear trend between standardized intermuscular fat* (IMF) and standardized digit symbol substitution test (DSST), Health ABC.**

|  | **DSST**** | | |
| --- | --- | --- | --- |
|  | **Beta** | **95% CI** | **P** |
| **IMF** | -0.08 | (-0.12, -0.03) | <0.001 |

**Adjusted for age, sex, race

**Supplemental Table 2. Metabolites with statistically significant associations with both intermuscular fat (IMF) and digit symbol substitution test (DSST), organized by class**

| **Metabolites associated with lower IMF and higher DSST scores** | | | | | | | | |
| --- | --- | --- | --- | --- | --- | --- | --- | --- |
|  | | **IMF** | | | | **DSST** | | |
| **Superclass/ Metabolites** | **Class/ *Subclass*** | **Beta** | **95% CI** | **FDR P** | **Beta** | | **95% CI** | **FDR P** |
| **Lipids and lipid-like molecules** |  |  |  |  |  | |  |  |
| LysoPC(22:6(4Z,7Z,10Z,13Z,16Z,19Z)) (HILICP) | Glycerophospholipids/ *Glycerophosphocholines* | -1.11 | (-1.59, -0.63) | 5.18E-05 | 0.76 | | (0.26, 1.26) | 0.02 |
| LysoPE(22:6(4Z,7Z,10Z,13Z,16Z,19Z)/0:0) | Glycerophospholipids / *Glycerophosphoethanolamines* | -0.71 | (-1.20, -0.22) | 0.01 | 0.74 | | (0.23, 1.25) | 0.02 |
| PC(22:6(4Z,7Z,10Z,13Z,16Z,19Z)/18:3(6Z,9Z,12Z)) | Glycerophospholipids / *Glycerophosphocholines* | -0.60 | (-1.10, -0.10) | 0.04 | 1.21 | | (0.69, 1.73) | 1.21E-04 |
| PE(16:0/18:0) | Glycerophospholipids / *Glycerophosphoethanolamines* | -0.60 | (-1.11, -0.09) | 0.05 | 1.10 | | (0.57, 1.63) | 5.96E-04 |
| **Nucleosides, nucleotides, and analogues** |  |  |  |  |  | |  |  |
| GDP | Purine nucleosides / *Purine ribonucleotides* | -0.69 | (-1.17, -0.20) | 0.02 | 0.72 | | (0.21, 1.22) | 0.03 |
| ADP | Purine nucleosides / *Purine ribonucleotides* | -0.74 | (-1.23, -0.26) | 8.67E-03 | 0.85 | | (0.35, 1.35) | 6.07E-03 |
| Uridine | Pyrimidine nucleosides / *Not Available* | -0.67 | (-1.16, -0.18) | 0.02 | 1.22 | | (0.72, 1.72) | 6.92E-05 |
| **Organic acids and derivatives** |  |  |  |  |  | |  |  |
| N-Methyl-proline | Carboxylic acids and derivatives / *Amino acids, peptides, and analogues* | -0.65 | (-1.14, -0.16) | 0.02 | 1.35 | | (0.84, 1.85) | 8.58E-06 |
| **Organic oxygen compounds** |  |  |  |  |  | |  |  |
| Glyceric acid | Organooxygen compounds / *Carbohydrates and carbohydrate conjugates* | -0.60 | (-1.09, -0.12) | 0.04 | 1.38 | | (0.87, 1.88) | 5.91E-06 |
| **Organoheterocyclic compounds** |  |  |  |  |  | |  |  |
| γ-Carboxyethyl Hydroxychroman | Benzopyrans / *1-benzopyrans* | -0.62 | (-1.13, -0.11) | 0.04 | 0.80 | | (0.27, 1.33) | 0.02 |
| Thiamine | Diazines / *Pyrimidines and pyrimidine derivatives* | -0.63 | (-1.15, -0.12) | 0.04 | 1.11 | | (0.58, 1.64) | 5.96E-04 |
| 3-Indolepropionic acid | Indoles and derivatives / *Indolyl carboxylic acids and derivatives* | -1.02 | (-1.51, -0.53) | 2.25E-04 | 0.75 | | (0.24, 1.25) | 0.02 |
| Niacinamide | Pyridines and derivatives / *Pyridinecarboxylic acids and derivatives* | -0.71 | (-1.19, -0.22) | 0.01 | 0.96 | | (0.46, 1.46) | 1.90E-03 |

| **Metabolites associated with higher IMF and lower DSST scores** | | | | | | | | |
| --- | --- | --- | --- | --- | --- | --- | --- | --- |
|  | | **IMF** | | | | **DSST** | | |
| **Superclass/ Metabolites** | **Class/ *Subclass*** | **Beta** | **95% CI** | **FDR P** | **Beta** | | **95% CI** | **FDR P** |
| **Lipids and lipid-like molecules** |  |  |  |  |  | |  |  |
| 5-hydroxyeicosatetraenoic acid | Fatty Acyls / *Eicosanoids* | 1.16 | (0.66, 1.66) | 4.75E-05 | -0.68 | | (-1.20, -0.15) | 0.05 |
| 3-Hydroxybutyrylcarnitine | Fatty Acyls / *Fatty acid esters* | 1.73 | (1.25, 2.22) | 1.86E-10 | -1.02 | | (-1.53, -0.52) | 8.67E-04 |
| Hexanoylcarnitine (CAS: 6418-78-6) | Fatty Acyls / *Fatty acid esters* | 1.47 | (0.98, 1.96) | 1.07E-07 | -0.92 | | (-1.43, -0.41) | 3.31E-03 |
| L-Acetylcarnitine | Fatty Acyls / *Fatty acid esters* | 1.06 | (0.57, 1.54) | 1.15E-04 | -0.71 | | (-1.21, -0.20) | 0.03 |
| cis-5-Tetradecenoylcarnitine | Fatty Acyls / *Fatty acid esters* | 0.76 | (0.26, 1.26) | 8.95E-03 | -0.78 | | (-1.29, -0.26) | 0.02 |
| trans-2-Dodecenoylcarnitine | Fatty Acyls / *Fatty acid esters* | 0.63 | (0.12, 1.13) | 0.04 | -0.73 | | (-1.24, -0.21) | 0.03 |
| Palmitic acid | Fatty Acyls / *Fatty acids and conjugates* | 1.34 | (0.84, 1.83) | 2.12E-06 | -0.67 | | (-1.18, -0.15) | 0.05 |
| Oleic acid | Fatty Acyls / *Fatty acids and conjugates* | 1.31 | (0.81, 1.81) | 3.72E-06 | -0.78 | | (-1.29, -0.26) | 0.02 |
| 11Z,14Z-Eicosadienoic acid | Fatty Acyls / *Fatty acids and conjugates* | 1.11 | (0.62, 1.60) | 7.32E-05 | -0.69 | | (-1.20, -0.17) | 0.04 |
| Eicosenoic acid | Fatty Acyls / *Fatty acids and conjugates* | 1.06 | (0.57, 1.55) | 1.33E-04 | -0.77 | | (-1.27, -0.26) | 0.02 |
| 10-Nonadecenoic acid | Fatty Acyls / *Fatty acids and conjugates* | 1.03 | (0.54, 1.53) | 2.30E-04 | -0.83 | | (-1.34, -0.31) | 0.01 |
| 2-Hydroxy-3-methyl-pentanoic acid or 2-Hydroxyisocaproic acid | Fatty Acyls / *Fatty acids and conjugates* | 0.96 | (0.43, 1.48) | 1.47E-03 | -0.70 | | (-1.24, -0.16) | 0.05 |
| Stearic acid | Fatty Acyls / *Fatty acids and conjugates* | 0.93 | (0.45, 1.41) | 7.95E-04 | -0.68 | | (-1.19, -0.18) | 0.04 |
| LysoPC(22:5(7Z,10Z,13Z,16Z,19Z)) | Glycerophospholipids/ *Glycerophosphocholines* | 0.74 | (0.25, 1.24) | 0.01 | -1.68 | | (-2.19, -1.17) | 2.01E-08 |
| **Nucleosides, nucleotides, and analogues** |  |  |  |  |  | |  |  |
| 1-Methyladenosine | Purine nucleosides / Not Available | 1.96 | (1.47, 2.44) | 7.78E-13 | -0.66 | | (-1.17, -0.15) | 0.05 |
| N4-Acetylcytidine | Pyrimidine nucleosides / *Not Available* | 1.62 | (1.14, 2.10) | 2.70E-09 | -0.69 | | (-1.20, -0.19) | 0.03 |
| **Organic acids and derivatives** |  |  |  |  |  | |  |  |
| Cystine | Carboxylic acids and derivatives / *Amino acids, peptides, and analogues* | 1.25 | (0.77, 1.73) | 4.86E-06 | -0.81 | | (-1.31, -0.31) | 9.89E-03 |
| Hydroxyproline | Carboxylic acids and derivatives/ *Amino acids, peptides, and analogues* | 0.65 | (0.12, 1.18) | 0.04 | -1.31 | | (-1.86, -0.75) | 8.95E-05 |
| Citric acid/Isocitric acid | Carboxylic acids and derivatives / *Tricarboxylic acids and derivatives* | 0.65 | (0.15, 1.14) | 0.03 | -0.67 | | (-1.19, -0.16) | 0.05 |
| Lactic acid | Hydroxy acids and derivatives / *Alpha hydroxy acids and derivatives* | 1.17 | (0.68, 1.65) | 2.24E-05 | -0.67 | | (-1.17, -0.16) | 0.04 |
| 3-Hydroxyoctanoic acid | Hydroxy acids and derivatives / *Medium-chain hydroxy acids and derivatives* | 0.60 | (0.11, 1.08) | 0.04 | -0.92 | | (-1.43, -0.42) | 2.91E-03 |
| **Organic nitrogen compounds** |  |  |  |  |  | |  |  |
| 5-Acetylamino-6-amino-3-methyluracil | Organonitrogen compounds / *N-arylamides* | 0.77 | (0.27, 1.27) | 8.86E-03 | -0.70 | | (-1.23, -0.18) | 0.04 |
| **Organoheterocyclic compounds** |  |  |  |  |  | |  |  |
| 1,7-Dimethyluric acid (C18N) | Imidazopyrimidines/ *Purines and purine derivatives* | 1.28 | (0.79, 1.78) | 4.91E-06 | -0.72 | | (-1.24, -0.21) | 0.03 |
| 1,7-Dimethyluric acid (HILICP) | Imidazopyrimidines/ *Purines and purine derivatives* | 1.08 | (0.59, 1.57) | 1.15E-04 | -0.95 | | (-1.46, -0.44) | 2.62E-03 |
| Caffeine | Imidazopyrimidines / *Purines and purine derivatives* | 1.31 | (0.82, 1.81) | 3.18E-06 | -0.66 | | (-1.18, -0.15) | 0.05 |
| Acisoga | Pyrrolidines / *N-alkylpyrrolidines* | 1.05 | (0.56, 1.54) | 1.43E-04 | -0.70 | | (-1.20, -0.19) | 0.03 |

**Supplemental Table 2. Subset of metabolites, where higher levels were associated with less IMF, but worse DSST among Health ABC participants, organized by class.**

|  |  | Intermuscular Fat | |  | Modified Mini-Mental State Exam | | |
| --- | --- | --- | --- | --- | --- | --- | --- |
| Superclass/ Metabolites | Class/ *Subclass* | Beta | 95% CI | FDR P | Beta | 95% CI | FDR P |
| **Lipids and lipid-like molecules** |  |  |  |  |  |  |  |
| y2_C18n_Adrenic_acid | Fatty Acyls/ *Fatty acids and conjugates* | 1.89 | (1.12,2.67) | 6.33E-05 | -1.19 | (-1.92,-0.46) | 0.02 |
| y2_HILICp_CAR_4_0_OH__ | Fatty Acyls / *Fatty acid esters* | 1.59 | (0.84,2.35) | 6.05E-04 | -1.02 | (-1.72,-0.31) | 0.03 |
| y2_C8p_PC_P_38_5__PC_O_38_6_ | Glycerophospholipids / *Glycerophosphocholines* | -1 | (-1.78,-0.22) | 0.04 | 1.33 | (0.60,2.05) | 7.70E-03 |
| y2_HILICp_PC_P_38_6__PC_O_38_7_ | Glycerophospholipids / *Glycerophosphocholines* | -1.08 | (-1.85,-0.31) | 0.02 | 1.2 | (0.48,1.92) | 0.01 |
| y2_C8p_LPC_20_5_ | Glycerophospholipids/ Glycerophosphocholines | -1.53 | (-2.27,-0.80) | 6.76E-04 | 1.57 | (0.88,2.25) | 4.31E-04 |
| y2_C8p_LPC_22_6_ | Glycerophospholipids/ Glycerophosphocholines | -1.94 | (-2.64,-1.23) | 5.79E-06 | 1.08 | (0.41,1.75) | 0.02 |
| y2_C8p_LPC_22_5_ | Glycerophospholipids/ Glycerophosphocholines | 1.03 | (0.30,1.77) | 0.02 | -1.95 | (-2.63,-1.27) | 5.25E-06 |
| **Organic acids and derivatives** |  |  |  |  |  |  |  |
| Cystine | Carboxylic acids and derivatives / *Amino acids, peptides, and analogues* | 1.29 | (0.51,2.06) | 6.97E-03 | -1.11 | (-1.83,-0.38) | 0.03 |
| y2_HILICp_N_Methyl_proline | Carboxylic acids and derivatives / *Amino acids, peptides, and analogues* | -1.01 | (-1.75,-0.27) | 0.03 | 1.63 | (0.94,2.32) | 3.67E-04 |
| y2_C18n_alpha_N_Phenylacetylglut | Carboxylic acids and derivatives / *Amino acids, peptides, and analogues* | 1.22 | (0.52,1.92) | 4.67E-03 | -1.01 | (-1.67,-0.36) | 0.02 |
| y2_HILICp_alpha_N_Phenylacetylgl | Carboxylic acids and derivatives / *Amino acids, peptides, and analogues* | 1.5 | (0.78,2.22) | 7.00E-04 | -1.19 | (-1.87,-0.52) | 9.50E-03 |
| y2_HILICp_Tryptophan | Carboxylic acids and derivatives / *Amino acids, peptides, and analogues* | -1.41 | (-2.25,-0.57) | 6.13E-03 | 1.77 | (1.00,2.55) | 4.31E-04 |
| y2_HILICn_Fumaric_acid_or_Maleic | Carboxylic acids and derivatives / *Dicarboxylic acids and derivatives* | 1.35 | (0.60,2.10) | 3.75E-03 | -1.11 | (-1.82,-0.41) | 0.02 |
| **Organoheterocyclic compounds** |  |  |  |  |  |  |  |
| α-Carboxyethyl Hydroxychroman | Benzopyrans */ 1-benzopyrans* | -1.2 | (-1.98,-0.41) | 0.01 | 1.21 | (0.47,1.95) | 0.02 |

**Supplemental Table 3. Summary of combined mediation analysis of top 4 potential mediating metabolites of the relationship between IMF and DSST among Health ABC participants.**

| **Variable** | **DSST** |  |  | **Proportion attenuated (%)** |
| --- | --- | --- | --- | --- |
|  | **Beta** | **SE** | **P** |  |
| **Direct** |  | | |  |
| IMF | -0.054 | 0.021 | 0.012 | -- |
| **Indirect** |  | | |  |
| Maslinic acid | -0.010 | 0.003 | 0.001 | 11.0 |
| 1-MNA | -0.010 | 0.003 | 0.001 | 11.0 |
| LPC(20:5) | -0.007 | 0.002 | 0.004 | 7.7 |
| Adrenic acid | -0.010 | 0.003 | 0.003 | 11.0 |
| **Total indirect** | -0.037 | 0.006 | <0.001 | 40.7 |

*Adjusted for age, race, gender, and muscle area

There is an inverse association between IMF and DSST, specifically, the total effect of IMF on DSST is -0.091, meaning that for every 1 cm^2^ increase in IMF, DSST decreases by 0.091 points. 40.7% of this association is attenuated by the top 4 metabolites identified in single mediation analyses: maslinic acid (11%), 1-MNA (11%), LPC(20:5) (7%), and adrenic acid (11%), indicating that these 4 metabolites together explain a substantial portion of the association between IMF and DSST.

X=Intermuscular Fat

Y=Digit Symbol Substitution Test

M=Metabolites:
Maslinic Acid
1-MNA
LPC(20:5)
Adrenic Acid

**Supplemental Table 4. Functional Annotations of metabolites that were associated with less Intermuscular Fat and better Digit Symbol Substitution Test scores.**

| **Metabolite** | **Function in relation to skeletal muscle** | **Function in relation to CNS** |
| --- | --- | --- |
| Adrenic acid | Isoprostane derivatives, such as those from adrenic acid are associated with vasoconstriction, pathogenesis of atherosclerosis and CVD^1^ | Adrenic acid is an n-6 polyunsaturated fatty acid (PUFA) that can be a biomarker of oxidative stress, especially in neuronal disorders and diseases, as well as bioactivities contributing to signaling and regulatory events in vivo. Third most important in the brain and particularly in myelin lipids. Abundant in adrenal gland and kidney, formed from arachidonic acid by chain elongation. Adrenic acid releases dihomo-isoprostane derivatives that can be markers of neuronal damage and disease in the brain.^1^ |
| Maslinic acid | Positively associated with muscle strength. Mice exposed to MA show Increase skeletal muscle mass and strength in older adults, via PI3/AKT/mTOR signaling, as well as TNFα signaling via NF-κB, TGF-β signaling, higher gf1 expression, lower expressions of Atrogin-1, Murf1 and Tgfb.^2^ | Can ameliorate cognitive impairment induced by cholinergic blockade in mice. This may be due to increased mature brain‐derived neurotrophic factors at specific timepoints of enhanced tissue type plasminogen activator expression.^3^. |
| 1-Methyl  nicotinamide | A muscle signaling molecule, or myokine, it enhances the utilization of energy stores in response to low muscle energy availability.^4^  1-MNA supplementation significantly improved physical performance in a 6-min walk test and reduced the percentage of older adults with severe fatigue after COVID-19.^5^ | Main metabolite of the amide form of vitamin B3 (nicotinamide). 1-MNA inhibits neuroinflammatory response, glial cell activation, and neuronal apoptosis in the hippocampus and frontal cortex in mice with lipopolysaccharide induced cognitive deficits.^6^  Nicotinamide, after metabolic conversion in the brain to N-methylnicotinamide, can lead to a blockade of choline clearance from the brain.^7^ |
| Lyso  Phosphatidyl  cholines | Circulating LysoPC were inversely associated with sarcopenia in older men.^8^  In mice models, comprehensive lipidomic analyses revealed a robust positive correlation of skeletal muscle lyso-PC and maximal force-generating capacity.^9^ | mediates pericyte loss, BBB disruption, demyelination, and motor function defects, and increases neurotoxicity of amyloid β peptide oligomer formation and neurotoxic protein aggregation. Some studies reported that plasma LPC was decreased in patients with AD and the LPC-to-PC ratio was also decreased.^10^ |
| Tryptophan | It increased % lean mass and decreased % body fat in adult mice on a low protein diet. It also upregulated mTor/eif4/p70s6k pathway molecules in muscles.^11^ | Beneficial effect son sleep and cognition- Precursor of the neurotransmitter serotonin.^12^  (Note: upstream regulator of kynurenin metabolism, precursor for the synthesis of NAD, nicotinic acid.) |
| Serotonin | Same as tryptophan | Low serotonin associated with decreased cognition. Serotonin may be connected with cognition via gut-brain axis.^13^ |
| 4-Pyridoxic acid | Levels vary according to vitamin B6 intake. Note: it’s the ratio of 4-PA/pyridoxal 5'-phosphate (PLP) that indicates VItB6 levels . Not clear why it would be a mediator. | A metabolite of Vitamin B6, which is essential for central nervous function and neurogenesis due to coenzyme action of their phospholipid derivatives in the brain metabolism of glucose and neurotransmitters.^14^ B-vitamin supplementation was found to slow cerebral gray matter atrophy. ^15^ |
| Phytanic acid | induce the expression of UCP1, an important protein in human skeletal muscles, where it may stimulate energy utilization.^16^ | Impairs Ca2+ metabolism, increases oxidative stress and de-energizes mitochondria in the brain.^17^ |
| Bilirubin | Elevated by exercise training, elite athletes may have significantly higher levels. bilirubin's antioxidant properties in response to a large number of reactive oxygen species (ROS)^18^ | Has antioxidant and anti-inflammatory properties and prevents dopaminergic neuron death by acting on tumor necrosis factor-alpha, which may be protective against Parkinson’s disease.^19^ |
| α-Carboxyethyl Hydroxychroman, γ-Carboxyethyl Hydroxychroman | It is not a naturally occurring metabolite. It is only found in those individuals exposed to this compound or its derivatives (Vit E supplements) or smoking. Vit E supplements (or no smoking)🡪 higher levels. we found this metabolite associated with lower IMAT and higher DSST🡪 it’s possible it’s just a proxy of lower smoking or higher vit E supplements. Not quite a real mediator of the association | Neuroprotective effects of vitamin E have been proposed, including modulating neurodegenerative signaling cascades, glutamate-induced neuronal cell death, as well as anti-inflammatory properties. Previous research has reported consistently vitamin E deficiently with MCI/AD. Vitamin E’s antioxidative properties have been shown to prevent amyloid-beta associated reactive plaques (characteristic of AD) from forming.^20^ |
| Uracil | The levels of uracil DNA glycosylase (the enzyme that is involved in excision repair) can vary in response to resistance training.  So, presumably, better muscle: lower enzyme, higher uracil levels ? unclear how it could act as a mediator | Uracil is a component of RNA. Folate is required to convert deoxyuridine monophosphate (dUMP) to deoxythymidine triphosphate (dTTP) which is essential for proper DNA synthesis. Folate shortage can cause excessive incorporation of uracil into the genome, causing DNA damage consistently observed in neurodegenerative diseases. Base excision repair was found to be deficient in both MCI and AD brain tissue.^21^ |
| Cholesterol  ester | Low serum cholesterol levels were associated with muscle weakness. Plasma cholesterol was found to be positively correlated with muscle force in mice. ^22^ | a cholesterol fatty acid ester, preferred for transport in plasma and for storage. Amyloid β binds cellular prion proteins increasing cholesterol concentrations, which is controlled by the cholesterol ester cycle via cholesterol ester hydrolases. Esterification, as expected, reduces cholesterol concentrations and disperses the above complexes. Cholesterol ester hydrolase inhibitors protected neurons and inhibition of cholesterol esterification increased amyloid β damage.^23^ |

**Supplemental Table 5. Functional Annotations of metabolites that were associated with more Intermuscular Fat and worse Digit Symbol Substitution Test scores.**

| **Class/Metabolite** | **Function** |
| --- | --- |
| **Lipids and lipid-like molecules** |  |
| Maslinic acid | Maslinic acid has been reported to ameliorate cognitive impairment induced by cholinergic blockade in mice. This may be due to increased mature brain‐derived neurotrophic factors at specific timepoints of enhanced tissue type plasminogen activator expression.^3^ |
| Phosphotidylethanolamine | Second most abundant glycerophospholipid in eukaryotic cells. Is the precursor of phosphatidylcholine, substrate for posttranslational modifications, affects membrane topology, promotes cell and organelle membrane fusion, oxidative phosphorylation, mitochondrial biogenesis, and autophagy. Excess PE can affect equilibrium and accumulation of amyloid β across the cell membrane. ^24^ |
| Lysophosphatidylcholine | Lysophosphatidylcholine mediates pericyte loss, vascular barrier disruption, demyelination, and motor function defects, and increases neurotoxicity of amyloid β peptide oligomer formation and neurotoxic protein aggregation. Some studies reported that plasma LPC was decreased in patients with AD and the LPC-to-PC ratio was also decreased.^10^ |
| Cholesterol ester | a cholesterol fatty acid ester, preferred for transport in plasma and for storage. Amyloid β binds cellular prion proteins increasing cholesterol concentrations, which is controlled by the cholesterol ester cycle via cholesterol ester hydrolases. Esterification, as expected, reduces cholesterol concentrations and disperses the above complexes. Cholesterol ester hydrolase inhibitors protected neurons and inhibition of cholesterol esterification increased amyloid β damage.^23^ |
| Adrenic acid | Severe peroxidation of membrane phospholipids rich in adrenic acid chains causes formation of reactive free radicals and consequently ferroptotic cell death.^25^ |
| Phytanic acid | Impairs Ca2+ metabolism, increases oxidative stress and de-energizes mitochondria in the brain.^17^ |
| **Organic acids and derivatives** |  |
| Phenylacetylglutamine | Byproduct of protein degradation by gut bacteria. Phenylacetylglutamine is formed when gut bacteria shifts from carbohydrate fermentation to protein metabolism, which may be a key feature of Parkinson’s disease microbiota.^26^ |
| Tryptophan | Tryptophan is an essential amino acid and is a precursor for the synthesis of proteins, NAD, nicotinic acid, and serotonin. 95% of tryptophan is metabolized through the kynurenine pathway, the remaining 5% is metabolized through the methoxyindole pathway to generate serotonin, which can further become melatonin.^12^ |
| N1,N12-Diacetylspermine | Spermine and its acetylated derivative N1,N12-Diacetylspermine have been associated with various cancer diagnosis and stages, including lung, breast, liver, colorectal, and urogenital.^27^ |
| Fumaric acid | Positive partial correlations were found between fumarate and malate, which could indirectly reflect upregulated enzymes in dementia patients such as succinate dehydrogenase and malate dehydrogenase. Enzymes were generally found to be downregulated in the first half of the TCA cycle and upregulated in the second half. This was also independent of APOE-ε4 genotype. Fumarate was previously reported to be positively associated with frailty.^28^  Argininosuccinate lyase (ASL) cleaves argininosuccinic acid to produce arginine and fumarate in the urea cycle. ASL deficiency is known to be associated with neurocognitive deficiencies (ADHD, developmental delay, seizures, and learning disability).^29^ |
| Maleic acid | Transcriptome profiling of neuroblastoma genes exposed to maleic acid showed that majority of the differentially expressed genes were associated with DNA binding and metal ion binding. Intracellular calcium and thiol levels were also affected, as well as expression of related genes.^30^ |
| Malic acid | Increased malate concentrations were found to be negatively associated with longevity and positively associated with frailty.^31^ |
| cis-Aconitic acid | Found to be protective against arterial stiffness.^32^ Previously found to be associated with AD.^33^ |
| **Organic oxygen compounds** |  |
| Sucrose | Sucrose may have a direct effect on the brain from its glucose component and a peripheral effect from fructose, which is metabolized by the liver first. Brain sensitivity to short-term drops/rises in glucose shows effects on performance in intensive cognitive tasks.^34^ |
| Lactose | Contributes the largest amount of galactose in human diet from consumption of milk/dairy products. Lactase hydrolyzes the β-1 → 4 glycosidic bond between galactose and glucose, the molecule components of lactose.^35^ |
| Trehalose | Usually ingested in the form of mushrooms or yeast. Known autophagy inducer in humans combined with anti-inflammatory effect can reduce accumulation of neurotoxic aberrant or misfolded proteins. Prospectively could be applied to treat neurodegeneration including Alzheimer’s, Parkinson’s, and Huntington’s disease.^36^ |
| p-Cresol glucuronide | a metabolite of p-cresol, a uremic toxin formed by bacterial fermentation of proteins in the large intestine. P-cresol has been associated with both Parkinson’s disease and autism spectrum disorder. P-cresol is formed when gut bacteria shifts from carbohydrate fermentation to protein metabolism, which may be a key feature of Parkinson’s disease microbiota.^26^ |
| Glucuronic acid | UDP-glucuronosyltransferases formed from glucuronic acid protect the brain against harmful lipophilic substances by metabolizing them as hydrophilic glucuronides.^37^ |
| **Organoheterocyclic compounds** |  |
| Bilirubin | Has antioxidant and anti-inflammatory properties and prevents dopaminergic neuron death by acting on tumor necrosis factor-alpha, which may be protective against Parkinson’s disease.^19^ |
| α-Carboxyethyl Hydroxychroman | Urinary metabolites of vitamin E. Vitamin E is a major lipid-soluble antioxidant. Neuroprotective effects of vitamin E have been proposed, including modulating neurodegenerative signaling cascades, glutamate-induced neuronal cell death, as well as anti-inflammatory properties. Previous research has reported consistently vitamin E deficiently with MCI/AD. Vitamin E’s antioxidative properties have been shown to prevent amyloid-beta associated reactive plaques (characteristic of AD) from forming.^20^ |
| Uracil | Uracil is a component of RNA. Folate is required to convert deoxyuridine monophosphate (dUMP) to deoxythymidine triphosphate (dTTP) which is essential for proper DNA synthesis. Folate shortage can cause excessive incorporation of uracil into the genome, causing DNA damage consistently observed in neurodegenerative diseases. Base excision repair was found to be deficient in both MCI and AD brain tissue.^21^ |
| 1-Methylnicotinamide | Main metabolite of the amide form of vitamin B3 (nicotinamide). 1-Methylnicotinamide (MNA) was able to inhibit neuroinflammatory response, glial cell activation, and neuronal apoptosis in the hippocampus and frontal cortex in mice with lipopolysaccharide induced cognitive deficits.^6^ |
| 1-Methyluric acid | Free radical scavenging function good for protecting against peroxyl oxidation.^38^ |
| 4-Pyridoxic acid | A metabolite of Vitamin B6, which is essential for central nervous function and neurogenesis due to coenzyme action of their phospholipid derivatives in the brain metabolism of glucose and neurotransmitters.^14^ B-vitamin supplementation was found to slow cerebral gray matter atrophy. ^15^ |
| Serotonin | Low serotonin associated with decreased cognition. Serotonin may be connected with cognition via gut-brain axis.^13^ |

**Supplemental Table 6. Metabolites with statistically significant associations with both intermuscular fat (IMF) and digit symbol substitution test (DSST) among White participants**

| **Metabolites** | | | | | | | |
| --- | --- | --- | --- | --- | --- | --- | --- |
|  | | **IMF** | | | **DSST** | | |
| **Superclass/ Metabolites** | **Class/ *Subclass*** | **Beta** | **95% CI** | **FDR P** | **Beta** | **95% CI** | **FDR P** |
| **Lipids and lipid-like molecules** |  |  |  |  |  |  |  |
| 15S-HETE | Fatty Acyls/*Eicosanoids* | 1.36 | (0.82,1.90) | 1.38E-05 | -1.06 | (-1.68,-0.45) | 6.91E-03 |
| 5-HETE | Fatty Acyls/Eicosanoids | 1.30 | (0.76,1.83) | 3.32E-05 | -1.18 | (-1.79,-0.57) | 2.02E-03 |
| Palmitoleic Acid | Fatty Acyls / *Fatty acids and conjugates* | 1.25 | (0.68,1.83) | 1.67E-04 | -0.98 | (-1.63,-0.33) | 0.022 |
| 10-Heptadecenoic acid | Fatty Acyls / *Fatty acids and conjugates* | 1.44 | (0.88,2.00) | 9.82E-06 | -1.16 | (-1.80,-0.52) | 4.46E-03 |
| 10-Nonadecenoic acid | Fatty Acyls / *Fatty acids and conjugates* | 0.86 | (0.31,1.41) | 7.63E-03 | -1.30 | (-1.92,-0.67) | 9.81E-04 |
| 11Z,14Z-Eicosadienoic acid | Fatty Acyls / *Fatty acids and conjugates* | 0.84 | (0.31,1.36) | 6.98E-03 | -1.02 | (-1.62,-0.42) | 7.54E-03 |
| Adrenic acid | Fatty Acyls / *Fatty acids and conjugates* | 1.52 | (1.00,2.05) | 5.09E-07 | -1.46 | (-2.06,-0.87) | 7.40E-05 |
| Eicosatrienoic acid | Fatty Acyls / *Fatty acids and conjugates* | 0.93 | (0.39,1.47) | 3.19E-03 | -1.01 | (-1.62,-0.40) | 9.33E-03 |
| Eicosenoic acid | Fatty Acyls / *Fatty acids and conjugates* | 0.69 | (0.15,1.23) | 0.03 | -0.97 | (-1.59,-0.36) | 0.01 |
| Oleic acid | Fatty Acyls / *Fatty acids and conjugates* | 1.25 | (0.72,1.78) | 5.54E-05 | -1.26 | (-1.86,-0.66) | 9.81E-04 |
| Palmitic acid | Fatty Acyls / *Fatty acids and conjugates* | 1.18 | (0.66,1.70) | 1.00E-04 | -1.06 | (-1.66,-0.47) | 4.81E-03 |
| Stearic acid | Fatty Acyls / *Fatty acids and conjugates* | 0.78 | (0.27,1.30) | 0.01 | -0.97 | (-1.56,-0.38) | 0.01 |
| 3-Carboxy-4-methyl-5-propyl-2-furanpropanoic acid | Fatty Acyls / *Fatty acids and conjugates* | -0.74 | (-1.25,-0.23) | 0.01 | 1.90 | (1.32,2.48) | 8.63E-08 |
| Myristoleic acid | Fatty Acyls / *Fatty acids and conjugates* | 0.86 | (0.37,1.36) | 2.82E-03 | -1.25 | (-1.81,-0.68) | 4.38E-04 |
| CAR(12:1) | Fatty Acyls / *Fatty acid esters* | 0.85 | (0.32,1.39) | 6.65E-03 | -1.13 | (-1.73,-0.52) | 3.25E-03 |
| CAR(14:1) | Fatty Acyls / *Fatty acid esters* | 0.73 | (0.20,1.26) | 0.02 | -1.17 | (-1.77,-0.57) | 2.02E-03 |
| CAR(2:0) | Fatty Acyls / *Fatty acid esters* | 1.03 | (0.51,1.54) | 6.27E-04 | -1.08 | (-1.66,-0.49) | 3.63E-03 |
| CAR(4:0(OH)) | Fatty Acyls / *Fatty acid esters* | 1.71 | (1.18,2.23) | 1.71E-08 | -1.57 | (-2.17,-0.97) | 2.14E-05 |
| Linoleic acid | Fatty Acyls/ *Lineolic acids and derivatives* | 0.91 | (0.37,1.45) | 4.07E-03 | -1.08 | (-1.70,-0.46) | 5.97E-03 |
| PC(38:6) | Glycerophospholipids / *Glycerophosphocholines* | -1.00 | (-1.49,-0.50) | 6.16E-04 | 1.61 | (1.05,2.18) | 3.86E-06 |
| PC(40:6) | Glycerophospholipids / *Glycerophosphocholines* | -0.86 | (-1.37,-0.35) | 4.11E-03 | 1.27 | (0.69,1.86) | 5.64E-04 |
| PC(40:9) | Glycerophospholipids / *Glycerophosphocholines* | -1.05 | (-1.54,-0.55) | 2.72E-04 | 1.64 | (1.08,2.19) | 3.26E-06 |
| PC(P-38:5)/PC(O-38:6) | Glycerophospholipids / *Glycerophosphocholines* | -0.92 | (-1.44,-0.39) | 2.78E-03 | 1.38 | (0.78,1.97) | 2.06E-04 |
| PC(P-38:6)/PC(O-38:7) | Glycerophospholipids / *Glycerophosphocholines* | -1.05 | (-1.57,-0.54) | 4.13E-04 | 1.22 | (0.64,1.80) | 9.42E-04 |
| LPC(22:6) | Glycerophospholipids / *Glycerophosphocholines* | -1.54 | (-2.03,-1.06) | 4.59E-08 | 1.17 | (0.61,1.73) | 9.61E-04 |
| LPC(16:0) | Glycerophospholipids / *Glycerophosphocholines* | -0.81 | (-1.35,-0.27) | 0.01 | 0.85 | (0.24,1.46) | 0.04 |
| LPC(20:5) | Glycerophospholipids / *Glycerophosphocholines* | -1.27 | (-1.76,-0.78) | 7.66E-06 | 1.61 | (1.05,2.16) | 3.26E-06 |
| LPC(22:5) | Glycerophospholipids / *Glycerophosphocholines* | 0.92 | (0.42,1.42) | 1.70E-03 | -1.47 | (-2.03,-0.90) | 2.14E-05 |
| LPC(22:6) | Glycerophospholipids / *Glycerophosphocholines* | -1.80 | (-2.28,-1.33) | 4.69E-11 | 1.46 | (0.92,2.01) | 1.48E-05 |
| LPC(P-16:0)/LPC(O-16:1) | Glycerophospholipids / *Glycerophosphocholines* | -0.68 | (-1.20,-0.17) | 0.03 | 0.81 | (0.22,1.39) | 0.04 |
| LPE(16:0) (C8p) | Glycerophospholipids/ *Glycerophosphoethanolamines* | -0.99 | (-1.53,-0.44) | 2.19E-03 | 1.27 | (0.64,1.89) | 1.37E-03 |
| LPE(20:0) | Glycerophospholipids/ *Glycerophosphoethanolamines* | -1.15 | (-1.67,-0.64) | 1.13E-04 | 0.78 | (0.19,1.36) | 0.05 |
| LPE(22:6) (C8p) | Glycerophospholipids/ *Glycerophosphoethanolamines* | -1.15 | (-1.65,-0.66) | 6.19E-05 | 1.22 | (0.66,1.79) | 5.64E-04 |
| LPE(16:0) (HILICp) | Glycerophospholipids/ *Glycerophosphoethanolamines* | -0.86 | (-1.40,-0.31) | 7.62E-03 | 0.86 | (0.23,1.48) | 0.04 |
| LPE(22:6) (HILICp) | Glycerophospholipids/ *Glycerophosphoethanolamines* | -0.90 | (-1.39,-0.40) | 2.09E-03 | 0.84 | (0.27,1.40) | 0.02 |
| PE(P-36:0)/PE(O-36:1) | Glycerophospholipids/ *Glycerophosphoethanolamines* | -0.63 | (-1.12,-0.14) | 0.03 | 0.89 | (0.33,1.45) | 0.01 |
| PE(P-38:6)/PE(O-38:7) | Glycerophospholipids/ *Glycerophosphoethanolamines* | -0.62 | (-1.14,-0.11) | 0.04 | 1.18 | (0.59,1.76) | 1.42E-03 |
| TG(56:10) | Glycerolipids / *Triradylcglycerols* | -0.60 | (-1.11,-0.09) | 0.05 | 1.35 | (0.78,1.93) | 1.71E-04 |
| TG(56:8) | Glycerolipids / *Triradylcglycerols* | -0.58 | (-1.07,-0.09) | 0.05 | 1.31 | (0.75,1.86) | 1.77E-04 |
| TG(58:10) | Glycerolipids / *Triradylcglycerols* | -0.70 | (-1.20,-0.20) | 0.02 | 1.42 | (0.85,1.99) | 6.21E-05 |
| TG(58:11) | Glycerolipids / *Triradylcglycerols* | -0.69 | (-1.19,-0.19) | 0.02 | 1.52 | (0.95,2.08) | 1.48E-05 |
| TG(58:9) | Glycerolipids / *Triradylcglycerols* | -0.98 | (-1.45,-0.50) | 4.64E-04 | 1.44 | (0.90,1.98) | 1.62E-05 |
| TG(60:12) | Glycerolipids / *Triradylcglycerols* | -0.72 | (-1.19,-0.24) | 0.01 | 1.46 | (0.91,2.00) | 1.48E-05 |
| Maslinic acid | Prenol lipids/ *Triterpenoids* | -1.14 | (-1.63,-0.66) | 5.26E-05 | 1.02 | (0.47,1.57) | 3.41E-03 |
| CE(14:0) | Steroids and steroid derivatives / *Steroid esters* | -0.75 | (-1.27,-0.22) | 0.02 | 0.83 | (0.23,1.43) | 0.04 |
| CE(20:5) | Steroids and steroid derivatives / *Steroid esters* | -0.77 | (-1.25,-0.29) | 6.65E-03 | 1.18 | (0.64,1.73) | 5.77E-04 |
| CE(22:6) | Steroids and steroid derivatives / *Steroid esters* | -1.15 | (-1.65,-0.65) | 9.31E-05 | 0.97 | (0.40,1.54) | 7.54E-03 |
| **Nucleosides, nucleotides, and analogues** |  |  |  |  |  |  |  |
| 1-Methyladenosine | Purine nucleosides / Not Available | 1.97 | (1.46,2.48) | 2.92E-11 | -1.00 | (-1.59,-0.41) | 7.54E-03 |
| Uridine | Pyrimidine nucleosides / *Not Available* | -0.81 | (-1.35,-0.27) | 0.010588 | 1.14 | (0.53,1.75) | 3.25E-03 |
| N4-Acetylcytidine | Pyrimidine nucleosides / *Not Available* | 1.65 | (1.12,2.17) | 5.77E-08 | -1.04 | (-1.64,-0.43) | 7.30E-03 |
| **Organic acids and derivatives** |  |  |  |  |  |  |  |
| Cystine | Carboxylic acids and derivatives / *Amino acids, peptides, and analogues* | 0.92 | (0.41,1.42) | 2.08E-03 | -0.80 | (-1.37,-0.22) | 0.04 |
| N-Formylmethionine | Carboxylic acids and derivatives / *Amino acids, peptides, and analogues* | 1.94 | (1.36,2.53) | 7.11E-09 | -0.95 | (-1.62,-0.28) | 0.03 |
| N-Acetylglutamic acid | Carboxylic acids and derivatives / *Amino acids, peptides, and analogues* | 1.86 | (1.32,2.40) | 2.88E-09 | -0.93 | (-1.56,-0.30) | 0.02 |
| Phenylacetylglutamine (C18N) | Carboxylic acids and derivatives / *Amino acids, peptides, and analogues* | 0.93 | (0.42,1.44) | 1.95E-03 | -0.81 | (-1.39,-0.23) | 0.04 |
| Phenylacetylglutamine (HILICp) | Carboxylic acids and derivatives / *Amino acids, peptides, and analogues* | 0.94 | (0.43,1.45) | 1.69E-03 | -0.85 | (-1.43,-0.27) | 0.02 |
| Tryptophan | Carboxylic acids and derivatives / *Amino acids, peptides, and analogues* | -1.15 | (-1.72,-0.59) | 4.36E-04 | 1.11 | (0.48,1.75) | 6.27E-03 |
| N-Methyl-proline | Carboxylic acids and derivatives / *Amino acids, peptides, and analogues* | -0.85 | (-1.39,-0.31) | 7.13E-03 | 1.23 | (0.62,1.84) | 1.38E-03 |
| N1,N12-Diacetylspermine | Carboximidic acids and derivatives / *Carboximidic acids* | 1.46 | (0.92,1.99) | 2.78E-06 | -1.29 | (-1.90,-0.69) | 7.41E-04 |
| Fumaric acid/Maleic acid | Carboxylic acids and derivatives / *Dicarboxylic acids and derivatives* | 1.16 | (0.63,1.70) | 1.72E-04 | -1.15 | (-1.75,-0.54) | 2.85E-03 |
| cis-Aconitic_acid | Carboxylic acids and derivatives / *Tricarboxylic acids and derivatives* | 1.58 | (1.06,2.10) | 1.53E-07 | -1.07 | (-1.67,-0.47) | 4.81E-03 |
| Citric acid/Isocitric acid | Carboxylic acids and derivatives / *Tricarboxylic acids and derivatives* | 0.80 | (0.27,1.33) | 9.68E-03 | -0.89 | (-1.50,-0.29) | 0.02 |
| Malic acid | Hydroxy acids and derivatives / *Beta hydroxy acids and derivatives* | 1.2 | (0.66,1.73) | 1.23E-04 | -1.14 | (-1.75,-0.53) | 3.23E-03 |
| **Organic oxygen compounds** |  |  |  |  |  |  |  |
| P-Cresol glucuronide | Organooxygen compounds / *Carbohydrates and carbohydrate conjugates* | 0.75 | (0.24,1.26) | 0.01 | -1.19 | (-1.77,-0.62) | 1.06E-03 |
| Glyceric acid | Organooxygen compounds / *Carbohydrates and carbohydrate conjugates* | -0.75 | (-1.27,-0.23) | 0.02 | 0.95 | (0.36,1.55) | 0.01 |
| Sucrose/Lactose/Trehalose | Organooxygen compounds / *Carbohydrates and carbohydrate conjugates* | 0.81 | (0.27,1.36) | 0.01 | -1.17 | (-1.79,-0.55) | 2.80E-03 |
| Glucuronic acid | Organooxygen compounds / *Carbohydrates and carbohydrate conjugates* | 1.22 | (0.68,1.76) | 1.22E-04 | -1.15 | (-1.77,-0.53) | 3.25E-03 |
| **Organic nitrogen compounds** |  |  |  |  |  |  |  |
| 5-Acetylamino-6-amino-3-methyluracil | Organonitrogen compounds/ *N-arylamides* | 1.09 | (0.43,1.75) | 4.96E-03 | -1.12 | (-1.87,-0.36) | 0.02 |
| **Organoheterocyclic compounds** |  |  |  |  |  |  |  |
| Uracil | Diazines / *Pyrimidines and pyrimidine derivatives* | -0.87 | (-1.41,-0.33) | 6.22E-03 | 1.06 | (0.44,1.67) | 7.12E-03 |
| Caffeine | Imidazopyrimidines / *Purines and purine derivatives* | 1.23 | (0.64,1.81) | 3.74E-04 | -0.93 | (-1.60,-0.25) | 0.04 |
| 1-Methyluric acid | Imidazopyrimidines / *Purines and purine derivatives* | 1.02 | (0.45,1.59) | 2.27E-03 | -1.09 | (-1.74,-0.44) | 8.51E-03 |
| 1,7-Dimethyluric acid (HILICp) | Imidazopyrimidines / *Purines and purine derivatives* | 1.07 | (0.49,1.66) | 1.76E-03 | -1.14 | (-1.80,-0.47) | 7.34E-03 |
| 3-Methylxanthine | Imidazopyrimidines / *Purines and purine derivatives* | 1.02 | (0.41,1.62) | 4.20E-03 | -0.93 | (-1.62,-0.24) | 0.05 |
| 1,7-Dimethyluric acid (C18n) | Imidazopyrimidines / *Purines and purine derivatives* | 1.38 | (0.79,1.96) | 5.89E-05 | -1.02 | (-1.69,-0.35) | 0.02 |
| Theophylline | Imidazopyrimidines / *Purines and purine derivatives* | 0.83 | (0.30,1.36) | 7.83E-03 | -0.88 | (-1.48,-0.28) | 0.03 |
| 4-Pyridoxic acid | Pyridines and derivatives / *Pyridinecarboxylic acids and derivatives* | -0.63 | (-1.16,-0.10) | 0.05 | 1.01 | (0.41,1.61) | 0.008294 |
| 1-Methylnicotinamide | Pyridines and derivatives / *Pyridinecarboxylic acids and derivatives* | -1.04 | (-1.55,-0.53) | 5.16E-04 | 0.98 | (0.39,1.56) | 8.51E-03 |
| Bilirubin | Tetrapyrroles and derivatives/ *Bilirubins* | -0.73 | (-1.29,-0.17) | 0.03 | 1.22 | (0.59,1.86) | 2.31E-03 |

**Supplemental Table 7. Metabolites with statistically significant associations with both intermuscular fat (IMF) and digit symbol substitution test (DSST) among Black participants**

| **Metabolites** | | | | | | | |
| --- | --- | --- | --- | --- | --- | --- | --- |
|  | | **IMF** | | | **DSST** | | |
| **Superclass/ Metabolites** | **Class/ *Subclass*** | **Beta** | **95% CI** | **FDR P** | **Beta** | **95% CI** | **FDR P** |
| **Lipids and lipid-like molecules** |  |  |  |  |  |  |  |
| LPC(15:0) | Glycerophospholipids / *Glycerophosphocholines* | -2.09 | (-3.07,-1.11) | 1.46E-03 | 1.51 | (0.57,2.45) | 0.02 |
| SM(d18:1/20:0) | Sphingolipids/ *Phosphosphingolipids* | -1.55 | (-2.56,-0.55) | 0.022 | 1.79 | (0.84,2.74) | 5.13E-03 |
| **Nucleosides, nucleotides, and analogues** |  |  |  |  |  |  |  |
| GDP | Purine nucleosides / Purine ribonucleotides | -1.36 | (-2.33,-0.39) | 0.036 | 1.47 | (0.56,2.39) | 0.02 |
| **Organoheterocyclic compounds** |  |  |  |  |  |  |  |
| 1-Methylnicotinamide | Pyridines and derivatives / *Pyridinecarboxylic acids and derivatives* | -1.80 | (-2.84,-0.76) | 0.010 | 2.18 | (1.20,3.16) | 6.08E-04 |
| Serotonin | Indoles and derivatives / *Tryptamines and derivatives* | -1.79 | (-2.82,-0.76) | 0.010 | 1.49 | (0.50,2.47) | 0.03 |

**Supplemental Table 8. Metabolites with statistically significant associations with both intermuscular fat (IMF) and digit symbol substitution test (DSST) among male participants**

| **Metabolites** | | | | | | | |
| --- | --- | --- | --- | --- | --- | --- | --- |
|  | | **IMF** | | | **DSST** | | |
| **Superclass/ Metabolites** | **Class/ *Subclass*** | **Beta** | **95% CI** | **FDR P** | **Beta** | **95% CI** | **FDR P** |
| **Lipids and lipid-like molecules** |  |  |  |  |  |  |  |
| Adrenic acid | *Fatty Acyls/ Fatty acids and conjugates* | 1.89 | (1.12,2.67) | 6.33E-05 | -1.19 | (-1.92,-0.46) | 0.02 |
| CAR(4:0(OH)) | Fatty Acyls / *Fatty acid esters* | 1.59 | (0.84,2.35) | 6.05E-04 | -1.02 | (-1.72,-0.31) | 0.03 |
| PC(P-38:5)/PC(O-38:6) | Glycerophospholipids / *Glycerophosphocholines* | -1 | (-1.78,-0.22) | 0.04 | 1.33 | (0.60,2.05) | 7.70E-03 |
| PC(P-38:6)/PC(O-38:7) | Glycerophospholipids / *Glycerophosphocholines* | -1.08 | (-1.85,-0.31) | 0.02 | 1.2 | (0.48,1.92) | 0.01 |
| LPC(20:5) | Glycerophospholipids/ *Glycerophosphocholines* | -1.53 | (-2.27,-0.80) | 6.76E-04 | 1.57 | (0.88,2.25) | 4.31E-04 |
| LPC(22:6) | Glycerophospholipids/ *Glycerophosphocholines* | -1.94 | (-2.64,-1.23) | 5.79E-06 | 1.08 | (0.41,1.75) | 0.02 |
| LPC(22:5) | Glycerophospholipids/ *Glycerophosphocholines* | 1.03 | (0.30,1.77) | 0.02 | -1.95 | (-2.63,-1.27) | 5.25E-06 |
| Phytanic acid | Prenol lipids / *Diterpenoids* | -1.22 | (-1.98,-0.46) | 9.26E-03 | 1.09 | (0.38,1.81) | 0.02 |
| Maslinic acid | Prenol lipids/ *Triterpenoids* | -1.18 | (-1.94,-0.43) | 0.01 | 1.69 | (0.99,2.39) | 3.45E-04 |
| **Nucleosides, nucleotides, and analogues** |  |  |  |  |  |  |  |
| GDP | Purine nucleosides / Purine ribonucleotides | -1.36 | (-2.33,-0.39) | 0.036 | 1.47 | (0.56,2.39) | 0.02 |
| **Organic acids and derivatives** |  |  |  |  |  |  |  |
| Cystine | Carboxylic acids and derivatives / Amino acids, peptides, and analogues | 1.29 | (0.51,2.06) | 6.97E-03 | -1.11 | (-1.83,-0.38) | 0.03 |
| N-Methyl-proline | Carboxylic acids and derivatives / Amino acids, peptides, and analogues | -1.01 | (-1.75,-0.27) | 0.03 | 1.63 | (0.94,2.32) | 3.67E-04 |
| Phenylacetylglutamine (C18N) | Carboxylic acids and derivatives/ Amino acids, peptides, and analogues | 1.22 | (0.52,1.92) | 4.67E-03 | -1.01 | (-1.67,-0.36) | 0.02 |
| Phenylacetylglutamine (HILICp) | Carboxylic acids and derivatives/ Amino acids, peptides, and analogues | 1.5 | (0.78,2.22) | 7.00E-04 | -1.19 | (-1.87,-0.52) | 9.50E-03 |
| Tryptophan | Carboxylic acids and derivatives/ Amino acids, peptides, and analogues | -1.41 | (-2.25,-0.57) | 6.13E-03 | 1.77 | (1.00,2.55) | 4.31E-04 |
| Fumaric acid/Maleic acid | Carboxylic acids and derivatives / Dicarboxylic acids and derivatives | 1.35 | (0.60,2.10) | 3.75E-03 | -1.11 | (-1.82,-0.41) | 0.02 |
| Malic acid | Hydroxy acids and derivatives / *Beta hydroxy acids and derivatives* | 1.28 | (0.52,2.03) | 5.73E-03 | -1.19 | (-1.89,-0.48) | 0.01 |
| **Organic oxygen compounds** |  |  |  |  |  |  |  |
| P-Cresol glucuronide | Organooxygen compounds / *Carbohydrates and carbohydrate conjugates* | 1.16 | (0.42,1.90) | 0.01 | -1.22 | (-1.91,-0.53) | 9.50E-03 |
| Sucrose/Lactose/Trehalose | Organooxygen compounds / *Carbohydrates and carbohydrate conjugates* | 0.96 | (0.20,1.73) | 0.05 | -1.36 | (-2.07,-0.64) | 4.43E-03 |
| Glucuronic acid | Organooxygen compounds / *Carbohydrates and carbohydrate conjugates* | 1.25 | (0.45,2.04) | 0.01 | -1.25 | (-1.99,-0.51) | 0.01 |
| **Organoheterocyclic compounds** |  |  |  |  |  |  |  |
| 3-Indolepropionic acid | Indoles and derivatives / *Indolyl carboxylic acids and derivatives* | -0.98 | (-1.73,-0.23) | 0.04 | 1.06 | (0.36,1.76) | 0.03 |
| 1-Methylnicotinamide | Pyridines and derivatives / *Pyridinecarboxylic acids and derivatives* | -1.42 | (-2.16,-0.68) | 1.88E-03 | 1.51 | (0.82,2.20) | 8.16E-04 |
| Niacinamide | Pyridines and derivatives / *Pyridinecarboxylic acids and derivatives* | -0.94 | (-1.68,-0.21) | 0.04 | 0.98 | (0.30,1.67) | 0.04 |
| Bilirubin | Tetrapyrroles and derivatives/ *Bilirubins* | -1.26 | (-2.02,-0.49) | 7.63E-03 | 1.59 | (0.87,2.30) | 7.09E-04 |

**Supplemental Table 9. Metabolites with statistically significant associations with both intermuscular fat (IMF) and digit symbol substitution test (DSST) among female participants**

| **Metabolites** | | | | | | | |
| --- | --- | --- | --- | --- | --- | --- | --- |
|  | | **IMF** | | | **DSST** | | |
| **Superclass/ Metabolites** | **Class/ *Subclass*** | **Beta** | **95% CI** | **FDR P** | **Beta** | **95% CI** | **FDR P** |
| **Lipids and lipid-like molecules** |  |  |  |  |  |  |  |
| LPC(14:0) | Glycerophospholipids / *Glycerophosphocholines* | -0.87 | (-1.54,-0.20) | 0.05 | 1.39 | (0.60,2.19) | 0.04 |
| Maslinic acid | Prenol lipids/ *Triterpenoids* | -1.12 | (-1.75,-0.49) | 4.16E-03 | 1.89 | (1.14,2.64) | 4.67E-04 |
| **Nucleosides, nucleotides, and analogues** |  |  |  |  |  |  |  |
| Uridine | Pyrimidine nucleosides / *Not Available* | -1.19 | (-1.81,-0.56) | 2.11E-03 | 1.22 | (0.48,1.96) | 0.04 |
| **Organic acids and derivatives** |  |  |  |  |  |  |  |
| N1,N12-Diacetylspermine | Carboximidic acids and derivatives / *Carboximidic acids* | 1.33 | (0.74,1.92) | 2.73E-04 | -1.19 | (-1.89,-0.49) | 0.04 |
| Homocitrulline | Carboxylic acids and derivatives / *Amino acids, peptides, and analogues* | 0.79 | (0.20,1.38) | 0.04 | -1.19 | (-1.89,-0.50) | 0.04 |
| 4-Hydroxyproline | Carboxylic acids and derivatives / *Amino acids, peptides, and analogues* | 0.99 | (0.32,1.66) | 0.02 | -1.43 | (-2.22,-0.63) | 0.04 |
| **Organoheterocyclic compounds** |  |  |  |  |  |  |  |
| α-Carboxyethyl Hydroxychroman | Benzopyrans */ 1-benzopyrans* | -1.18 | (-1.82,-0.54) | 2.73E-03 | 1.33 | (0.57,2.09) | 0.04 |
| Thiamine | Diazines / *Pyrimidines and pyrimidine derivatives* | -1.03 | (-1.70,-0.36) | 0.01 | 1.50 | (0.70,2.29) | 0.03 |
| 1-Methylnicotinamide | Pyridines and derivatives / *Pyridinecarboxylic acids and derivatives* | -1.13 | (-1.77,-0.48) | 4.61E-03 | 1.26 | (0.49,2.02) | 0.05 |

**Supplemental Figure 1. Flowchart of Study Participants**

All enrolled Health ABC participants had cognition data (n=3030)

No data on thigh intermuscular fat (IMF)
(n=62)

Had data on both cognition and IMF (n=2968)

No data on metabolites

(n=580)

Has data on cognition, IMF, and metabolites (n=2388)

Total Health ABC participants (n=3075)

No cognition data
(n=45)

**Supplemental Figure 2. Pathway Enrichment Analysis for metabolites**

A. **Metabolites**

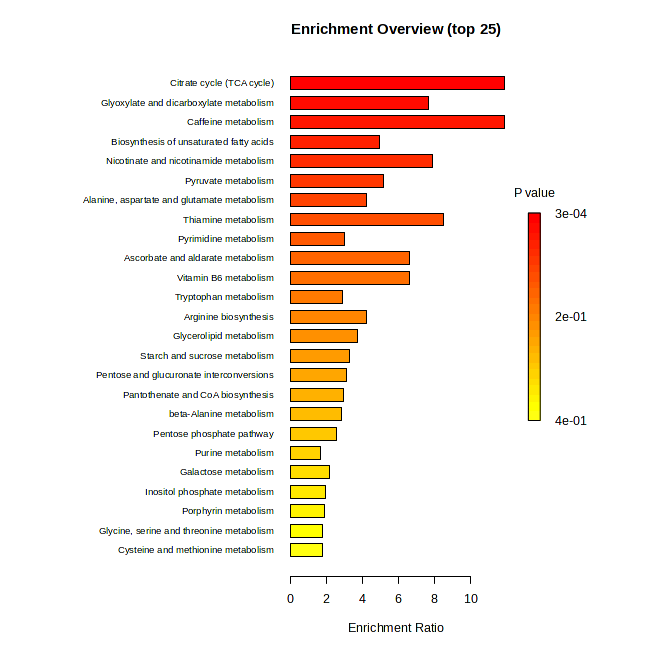

| **Pathway** | **Kegg Pathway Metabolite Count** | **Matched Metabolite Count** | **FDR P** | **Metabolites** |
| --- | --- | --- | --- | --- |
| Citrate cycle (TCA cycle) | 20 | 4 | 0.0207 | malic acid, cis-aconitic acid, citric acid, fumaric acid |

B. **Lipids**

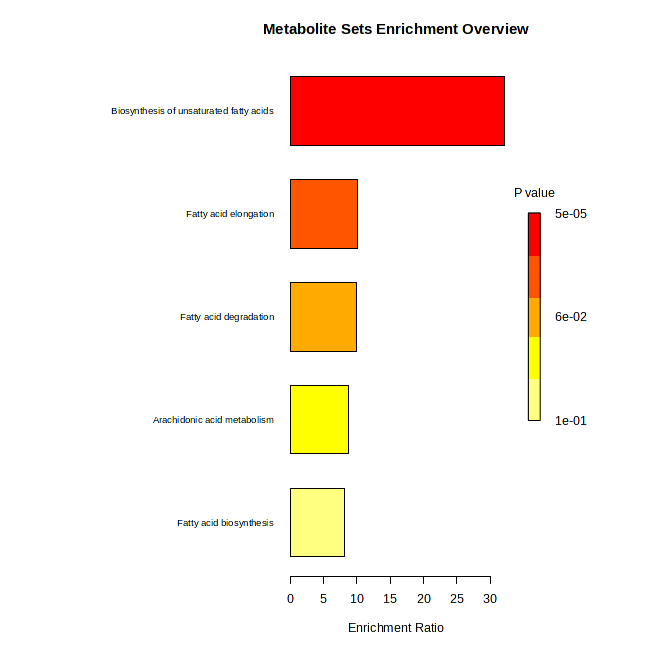

| **Pathway** | **Kegg Pathway Metabolite Count** | **Matched Metabolite Count** | **FDR P** | **Metabolites** |
| --- | --- | --- | --- | --- |
| Biosynthesis of unsaturated fatty acids | 36 | 3 | 0.00371 | palmitic acid, stearic acid, oleic acid |

**Supplemental Figure 3. Associations of top attenuating metabolites with IMF and DSST by race and sex**

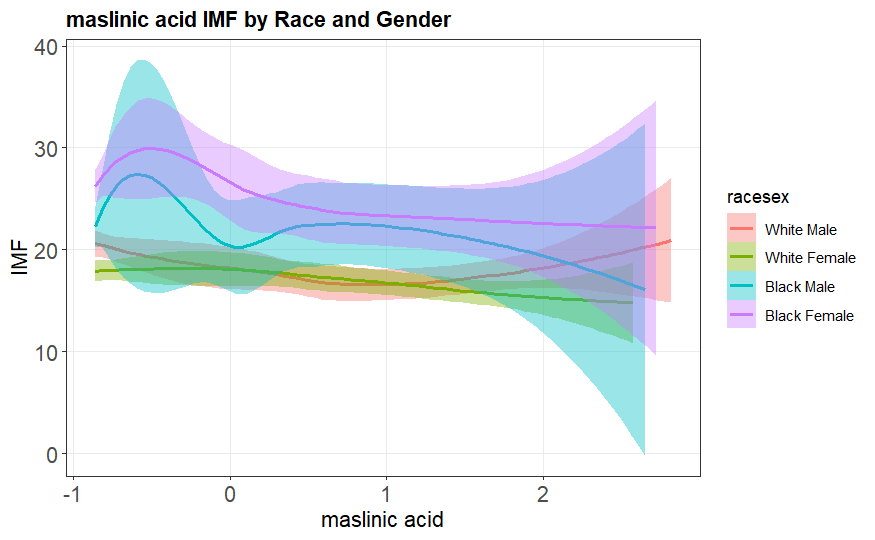

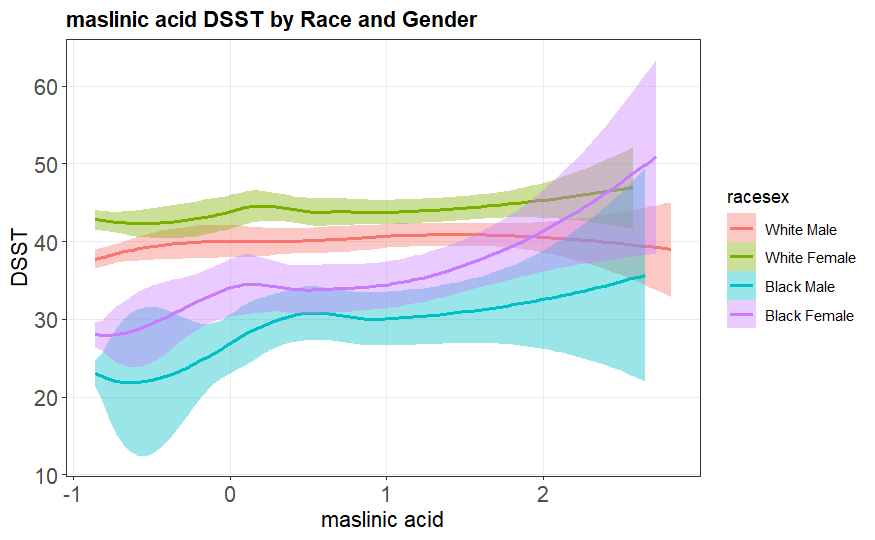

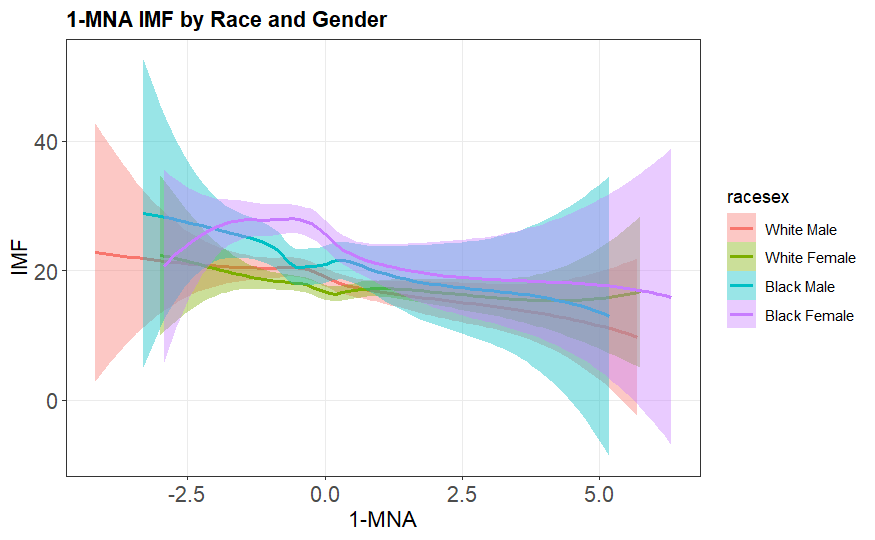

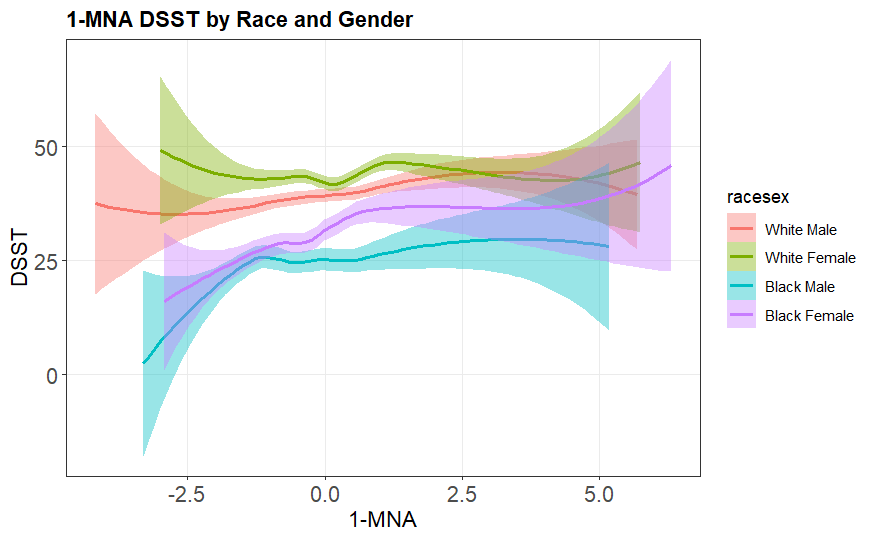

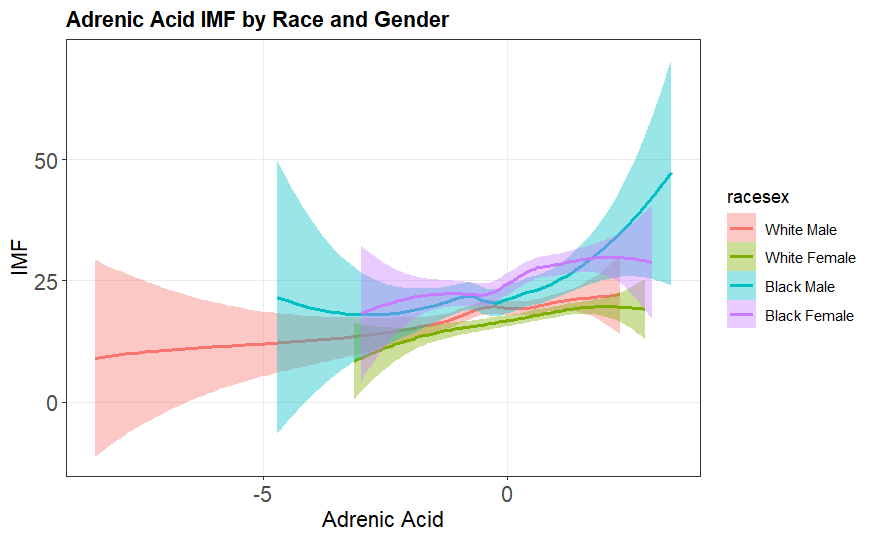

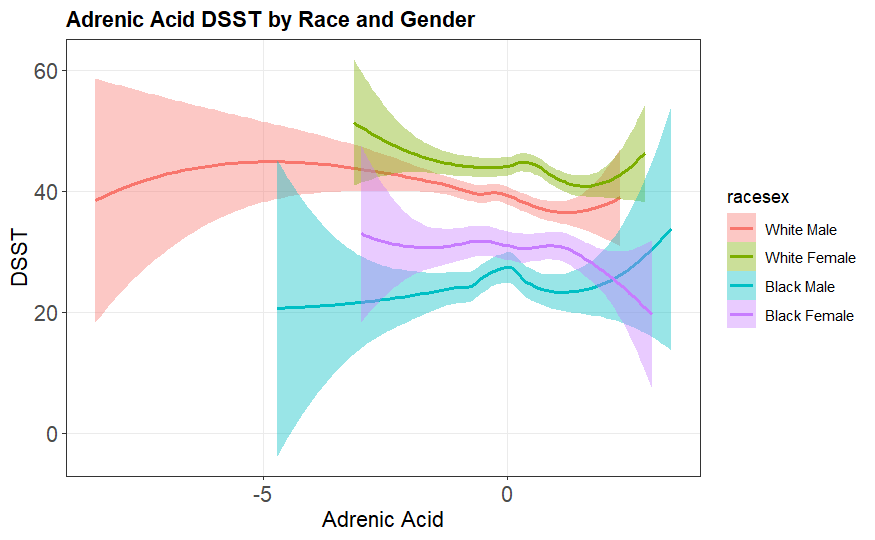

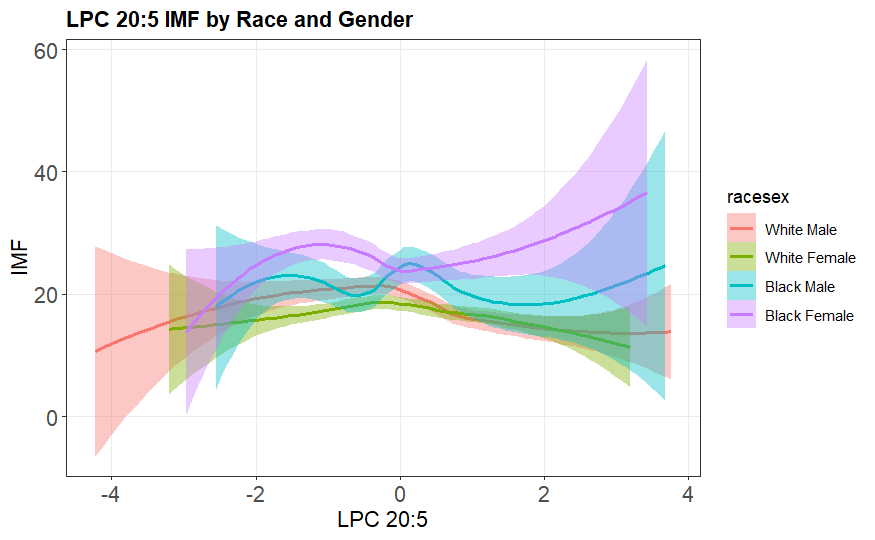

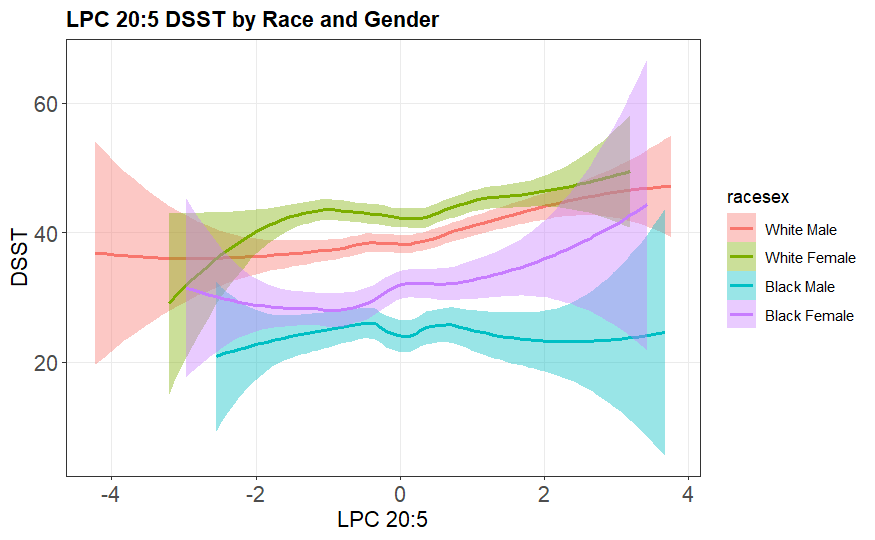

**Supplemental Figure 4. Pathway Enrichment Analysis for metabolites among Whites**

A. **Metabolites**

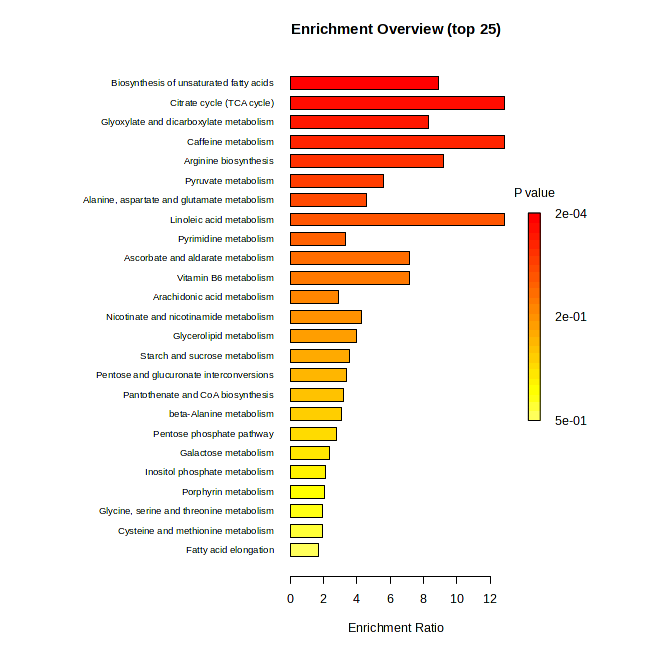

| **Pathway** | **Kegg Pathway Metabolite Count** | **Matched Metabolite Count** | **FDR P** | **Metabolites** |
| --- | --- | --- | --- | --- |
| Biosynthesis of unsaturated fatty acids | 36 | 5 | 0.00748 | palmitic acid, stearic acid, oleic acid, linoleic acid, dihomo-gamma-linolenic acid |
| Citrate cycle (TCA cycle) | 20 | 4 | 0.00748 | malic acid, cis-aconitic acid, citric acid, fumaric acid |
| Glyoxylate and dicarboxylate metabolism | 31 | 4 | 0.0288 | cis-aconitic acid, citric acid, malic acid, glyceric acid |

B. **Lipids**

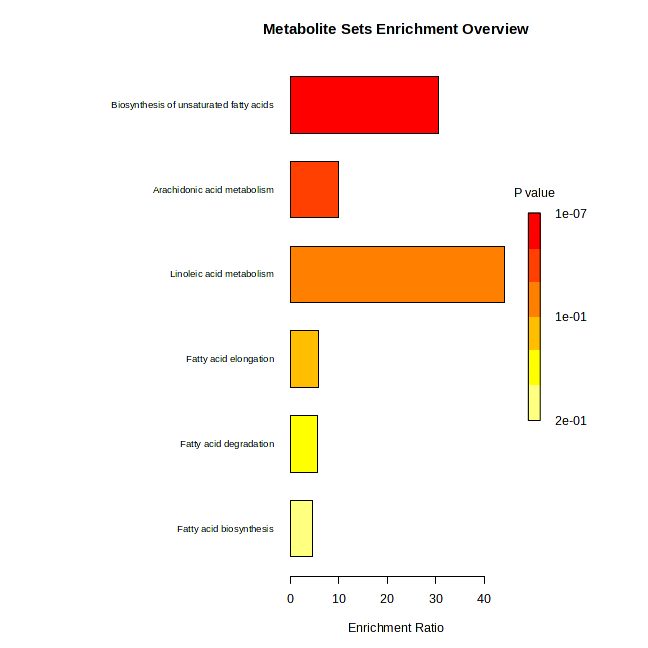

| **Pathway** | **Kegg Pathway Metabolite Count** | **Matched Metabolite Count** | **FDR P** | **Metabolites** |
| --- | --- | --- | --- | --- |
| Biosynthesis of unsaturated fatty acids | 36 | 5 | 8.56E-06 | palmitic acid, stearic acid, oleic acid, linoleic acid, dihomo-gamma-linoleic acid |

**Supplemental Figure 5. Pathway Enrichment Analysis for metabolites among Blacks**

A. **Metabolites**

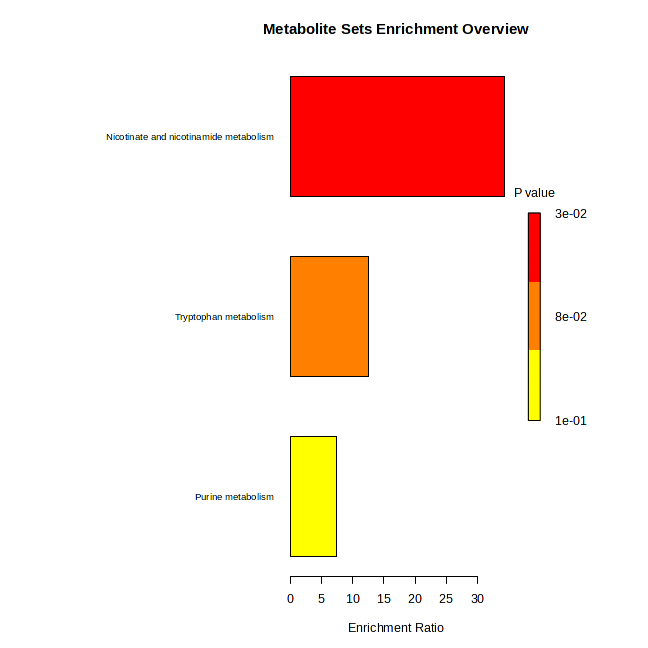

| **Pathway** | **Kegg Pathway Metabolite Count** | **Matched Metabolite Count** | **FDR P** | **Metabolites** |
| --- | --- | --- | --- | --- |
| Nicotinate and nicotinamide metabolism | 15 | 1 | 1 | 1-methyl nicotinamide |
| Tryptophan metabolism | 41 | 1 | 1 | serotonin |
| Purine metabolism | 70 | 1 | 1 | GDP |

**Supplemental Figure 6. Pathway Enrichment Analysis for metabolites among Males**

A. **Metabolites**

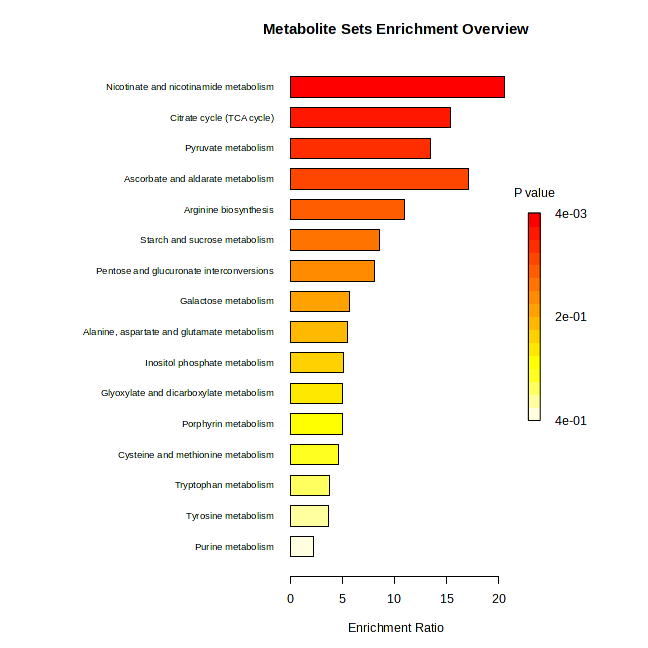

| **Pathway** | **Kegg Pathway Metabolite Count** | **Matched Metabolite Count** | **FDR P** | **Metabolites** |
| --- | --- | --- | --- | --- |
| Nicotinate and nicotinamide metabolism | 15 | 2 | 0.238 | niacinamide, 1-methylnicotinamde |
| Citrate cycle (TCA cycle) | 20 | 2 | 0.238 | malic acid, fumaric acid |
| Pyruvate metabolism | 23 | 2 | 0.238 | malic acid, fumaric acid |

**Supplemental Figure 7. Pathway Enrichment Analysis for metabolites among Females**

A. **Metabolites**

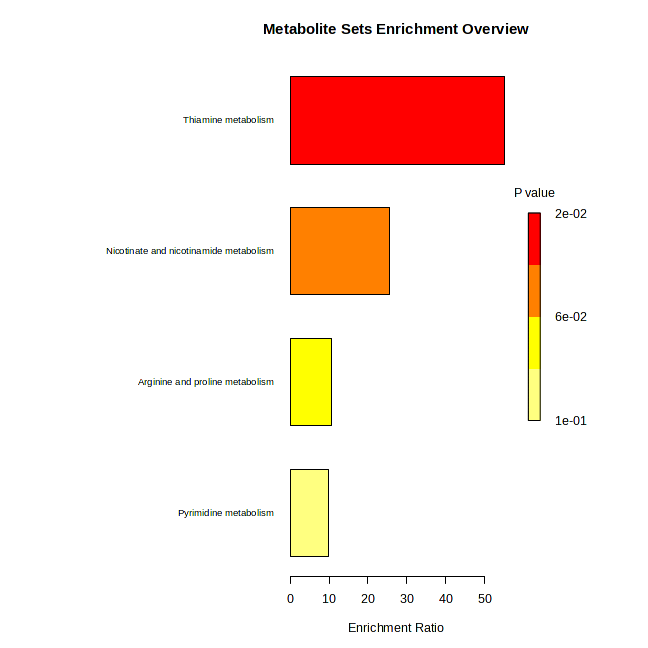

| **Pathway** | **Kegg Pathway Metabolite Count** | **Matched Metabolite Count** | **FDR P** | **Metabolites** |
| --- | --- | --- | --- | --- |
| Thiamine metabolism | 7 | 1 | 1 | thiamine |
| Nicotinate and nicotinamide metabolism | 15 | 1 | 1 | 1-methylnicotinamide |
| Arginine and proline metabolism | 36 | 1 | 1 | 4-hydroxyproline |
| Pyrimidine metabolism | 39 | 1 | 1 | uridine |
